# Model-driven mitigation measures for reopening schools during the COVID-19 pandemic

**DOI:** 10.1101/2021.01.22.21250282

**Authors:** Ryan S. McGee, Julian R. Homburger, Hannah E. Williams, Carl T. Bergstrom, Alicia Y. Zhou

## Abstract

Reopening schools is an urgent priority as the COVID-19 pandemic drags on. To explore the risks associated with returning to in-person learning and the value of mitigation measures, we developed stochastic, network-based models of SARS-CoV-2 transmission in primary and secondary schools. We find that a number of mitigation measures, alone or in concert, may reduce risk to acceptable levels. Student cohorting, in which students are divided into two separate populations that attend in-person classes on alternating schedules, can reduce both the likelihood and the size of outbreaks. Proactive testing of teachers and staff can help catch introductions early, before they spread widely through the school. In secondary schools, where the students are more susceptible to infection and have different patterns of social interaction, control is more difficult. Especially in these settings, planners should also consider testing students once or twice weekly. Vaccinating teachers and staff protects these individuals and may have a protective effect on students as well. Other mitigations, including mask-wearing, social distancing, and increased ventilation, remain a crucial component of any reopening plan.

## Introduction

As the COVID-19 pandemic accelerated in early 2020, schools around the world closed in an effort to preempt school-associated transmission and protect their students, teachers, and staff. By mid-April of that year, 195 countries had closed their schools in response to COVID-19, affecting more than 1.5 billion students.^1^ In the United States (US), schools were among the first organizations to close, and many remained closed or transitioned to remote learning through the end of the 2019-20 school year. Some remain closed today. While remote learning affords students the opportunity to continue their education, it fails to provide many of the crucial benefits students typically receive through in-person schooling.^2^ There is an urgent need to evaluate the effectiveness of evidence-based strategies that would allow children, teachers, and staff to safely return to in-person learning.

To date, widespread community transmission, conflicting public health guidance, and the emergence of new SARS-CoV-2 variants associated with higher transmissibility have compounded the challenges schools face when reopening.^3–5^ Numerous epidemiological models have been developed to forecast the spread of SARS-CoV-2 or compare the effectiveness of mitigation strategies in communities or large populations.^6–12^ However, only a few models have focused on the unique demographic and contact structures of primary and secondary school settings.^13–15^

Case studies suggest that primary schools have a lower risk of transmission compared to secondary schools ^16–19^. Two principal causes could be at play. First, younger children are less likely than adolescents or adults to become infected with SARS-CoV-2, ^20^ and less likely to experience symptomatic or severe disease.^21,22^ Second, primary and secondary schools have different contact structures. Primary school students have fewer contacts and typically spend the full day with a single teacher and the same group of students. By contrast, secondary school students move between classrooms and encounter multiple teachers and groups of students each day.

We have developed epidemiological models to simulate the spread of SARS-CoV-2 amongst students, teachers, and staff in both primary and secondary schools. Here, we use these models to better understand the risks of reopening schools and to explore the effectiveness of different mitigation strategies: cohorting students, proactive testing, quarantine protocols, and vaccinating teachers and staff.

## Model and methods

### A stochastic network-based model of SARS-CoV-2 transmission

We use the SEIRS+ modeling framework (https://github.com/ryansmcgee/seirsplus) to study the dynamics of disease transmission in school populations. SEIRS+ builds upon classic SEIR compartment models that divide the population into susceptible (*S*), exposed (*E*), infectious (*I*), and recovered (*R*) individuals and track the transitions of individuals among these states.^23^ The basic SEIR model is a deterministic model of a homogeneous population with well-mixed interactions. However, accounting for demographic heterogeneity and the structure of contact networks is particularly important when evaluating control strategies that perturb the contact network (e.g., social distancing) or make use of it (e.g., contact tracing).^24,25^ For disease control, modeling stochasticity is crucial to understand the distribution of potential outcomes, especially in smaller populations.

To incorporate these important aspects of disease dynamics, we use the SEIRS+ modeling framework to implement an extended SEIR model of SARS-CoV-2 transmission on stochastic dynamical networks. Individuals are represented as nodes in a contact network. Parameters, interactions, interventions, and residence times in each compartment are specified on an individual-by-individual basis. This allows us to model realistic heterogeneities in disease, transmission, and behavioral parameters—which are particularly important when considering SARS-CoV-2 transmission dynamics in small, age-stratified school populations. The disease dynamics are summarized in Figure 1 and described in detail in Appendix A.1. Parameter settings are outlined in Appendix A.2.

**Figure 1:**
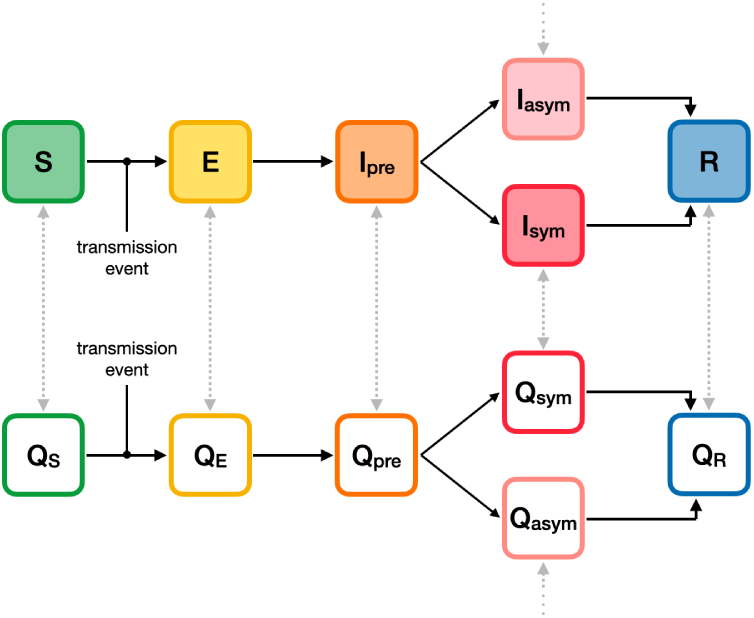
Compartment model. The progression of disease states in the Extended SEIR Network Model is represented by the compartments shown. Susceptible (*S*) individuals become infected (exposed) following transmissive contact with an infectious individual. Newly exposed (*E*) individuals undergo a latent period, during which time they are infected but not contagious. Infected individuals then progress to a pre-symptomatic infectious state (*I*_pre_), in which they are contagious but not yet presenting symptoms. Some infectious individuals go on to develop symptoms (*I*_sym_); while others will remain asymptomatic (*I*_asym_). At the conclusion of the infectious period, infected individuals enter the recovered state (*R*) and are no longer contagious or susceptible to infection. The unshaded compartments represent quarantined individuals in the respective disease states. Individuals are moved into a quarantine compartment upon isolation due to symptoms or a positive test (gray arrows).

We model infection as transmitted largely along a network of close contacts. Close contacts are individuals with whom one has repeated, sustained, or close-proximity interactions on a regular basis: classmates, friends, housemates, or other close relationships. Disease transmission can also occur among casual contacts—individuals who are not on one’s contact network, but with whom one has incidental, brief, or superficial interactions. A network locality parameter sets the relative frequency and weight of transmission among close and casual contacts (Appendix A.1.4). In both primary and secondary school settings, we assume that 80% of transmission occurs between close contacts specified by the networks.^26^ Exposure to the community is modeled by randomly introducing new cases to the school population at a rate that corresponds to the community prevalence—see the Community prevalence and case introduction rate section below.

The likelihood that a susceptible individual becomes infected depends on the prevalence of infectious individuals among their contacts, the transmissibilities of these contacts, and their own susceptibility to infection (Appendix A.1.4). Because individuals with large numbers of contacts—teachers, for example—are unlikely to interact as closely with each individual contact, we assume a logarithmic rather than linear scaling of transmission opportunity as a function of network degree (Appendix A.1.4.2). Independent of connectivity, we assume an over-dispersed distribution of individual variation in biological transmissibility (Appendix A.2.3), which corresponds to the observation that 80% of SARS-CoV-2 transmission may be attributable to 20% of infectious individuals.^27–29^ This distribution of individual transmissibilities is calibrated to a nominal basic reproduction number *R*_0_ for the population. While the *R*_0_ of SARS-CoV-2 varies, many estimates place *R*_0_ upward of 2.5-3.0 without intervention, depending on the variants that are present in a given population.^30–35^ As a baseline, we assume that schools will implement sufficient mitigation measures, such as mask wearing, physical distancing, and increased ventilation, to reduce *R*_0_ to 1.5 in the school population. More aggressive mitigation measures may bring the baseline *R*_0_ in schools closer to 1.0. Results for other values of *R*_0_ are discussed in Appendix B.

Individuals in any disease state may enter quarantine due to symptoms or in response to a positive test result. The effect of isolating individuals is modeled by introducing compartments that represent quarantined individuals who do not make transmissive contact with others outside of the home (Figure 1, Appendix A.2.4.3). Individuals remain in quarantine for 10 days, ^36^ at which time they transition to the non-quarantine compartment corresponding to their present disease state. We assume that 20% of symptomatic individuals self-isolate upon the onset of symptoms ^37^ (Appendix B.4). At baseline, only the symptomatic or positive individual is isolated, but we go on to consider scenarios where classroom contacts of positive students are also isolated (Appendix A.2.6.4).

### Model considerations for primary schools versus secondary schools

We use distinct models for primary and secondary schools, with different contact networks reflecting the social structures in each setting (Appendix A.2). We assume primary school children are 60% as susceptible as adults, while secondary school students are equally susceptible to adults.^16,20^ Our primary school model encompasses a school with 480 students, 24 teachers, and 24 additional staff. Primary school students have close contacts with their teacher, classmates, and other children in their household (e.g., siblings). For our secondary school model, we simulate a school with 800 students distributed across four grades, 125 teachers, and 75 additional staff. Secondary school students have close contacts with six teachers, with other students in their grade and social groups, and with other students in their households. Secondary school networks are parameterized such that connectivity statistics (e.g., mean degree, CV^2^ of degree, clustering coefficient) are in line with empirical studies of secondary school contact networks (^38–41^, Appendix A.2.4.2). Both settings feature a network of close contacts among teachers and staff. A new random network is generated for each simulation replicate. Example network diagrams for each school setting are shown in Figure 2. Detailed descriptions of the contact network structures and their generation are provided in Appendix A.2.4.

**Figure 2:**
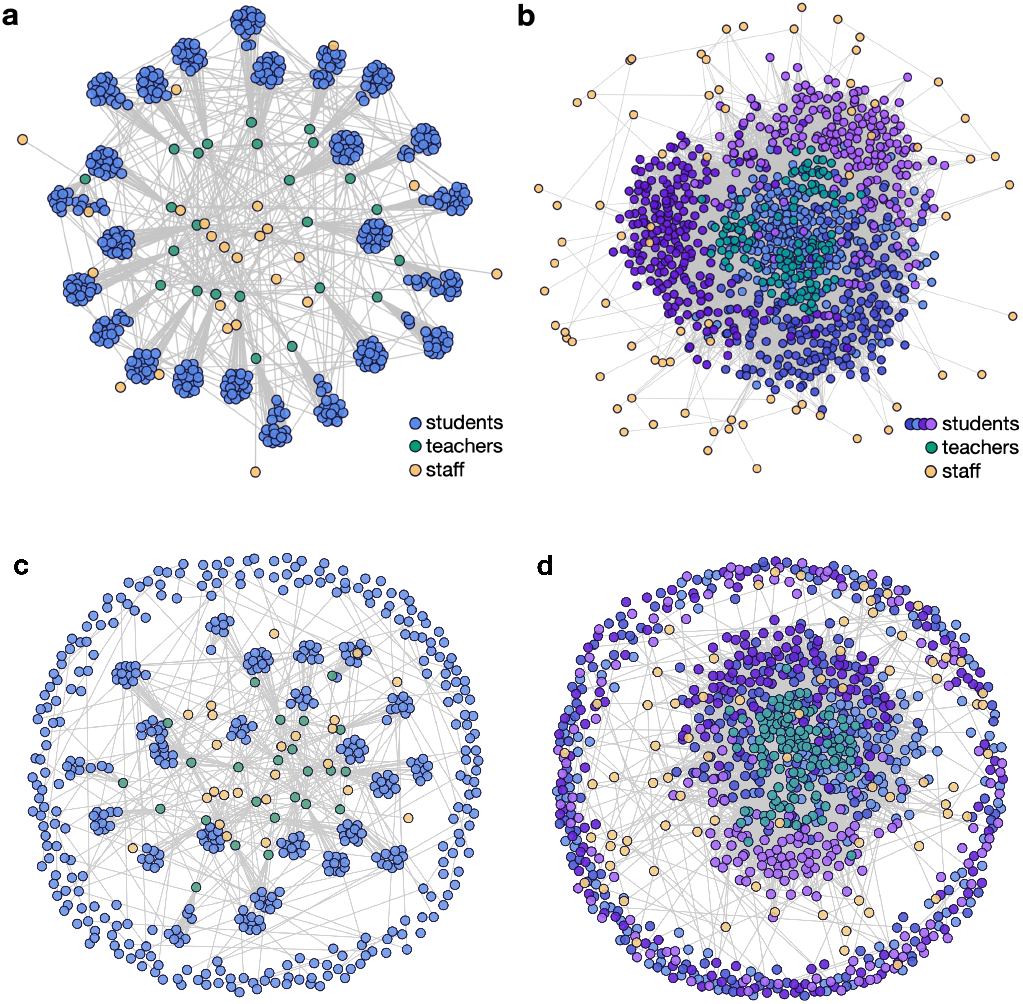
Network structures for primary and secondary schools. Each individual is represented by a circle, with grey lines connecting close contacts. (**a**) Primary school students (blue) are organized into classes with close contacts between all students in each classroom as well as a single teacher (green). School staff (yellow) interact with teachers and other staff. (**b**) Secondary school students (shades of blue and purple indicating grade levels) move between classrooms and have close contact with six teachers (green) each. School staff (yellow) interact with teachers and other staff. Secondary school students are clustered into loose social groups and are more likely to interact with other students in the same grade. (**c**,**d**) Example contact networks for primary and secondary schools, respectively, on a given day in a cohorting strategy in which students are divided into two groups that alternate in-person learning. Students that are in school on a given day (interior nodes) maintain the same school interactions as in the baseline networks. Students in the out-of-school cohort (peripheral nodes) make connections with any students that share their household (students in the same household are assigned to the same cohort in our model), but are disconnected from all other students and teachers. Students alternate between these interaction patterns according to a weekly or daily cohorting schedule.

### Community prevalence and case introduction rate

To account for the effect of community prevalence on COVID-19 dynamics in schools, we model scenarios in which new cases are introduced into the school population stochastically at rates corresponding to daily, weekly, or monthly introductions on average (Appendix A.2.5). When the effective community reproduction number *R*_eff_ is in the 1.0–2.0 range, these rates approximately correspond to the community prevalences shown in Table Appendix A.2.5. We also consider the consequences of a single introduction; in this scenario, all replicates start off with the case introduction occurring on the first day of the simulation.

### Simulations

To capture stochastic variability in outcomes, we report 1,000 replicates for each parameter set. Each replicate simulation tracks the progression of an outbreak that begins with the introduction of a single infected individual in an otherwise disease-free school population. The simulation begins on a random day of the week with the introduction of an initial case. Additional introductions may occur throughout the simulation at a Poisson rate reflecting the community prevalence. School is in session 5 days a week, and we assume that no close contacts are made outside of the household on weekends. Weekend transmission among casual contacts does occur. The simulation proceeds for 150 days to represent a school semester.

To allows comparisons across scenarios with different community prevalences, we report the percentage of cases attributable to transmissions within the school population (i.e., excluding introduced cases attributable to exogenous community exposure). These transmissions may occur either at school or among school-affiliated individuals while off campus, and are hereafter collectively described as “school transmission”. We define “sizable outbreaks”, as simulation runs where more than 5% of the population becomes infected in school over the course of the semester (150 days). While schools that experience sizable outbreaks are likely to stop in-person learning before very large case counts are realized, these data provide information about the probability of epidemic trajectories that could require such action.

## Results

### The effect of community prevalence

The prevalence of COVID-19 in the community impacts the risk of transmission in schools. Figure 3 shows the percentage of the school population infected in primary and secondary schools over the course of a semester when only basic mitigation strategies (e.g. distancing, hygiene, and mask wearing) are in place. Higher COVID-19 prevalence in the surrounding community increases the probability of a sizable outbreak in primary and secondary schools alike. When community prevalence is so high that new introductions occur on a daily basis (between 0.25–1.0%), our simulations suggest that even aggressive mitigation strategies cannot prevent sizable outbreaks (Figure 5).

**Figure 3:**
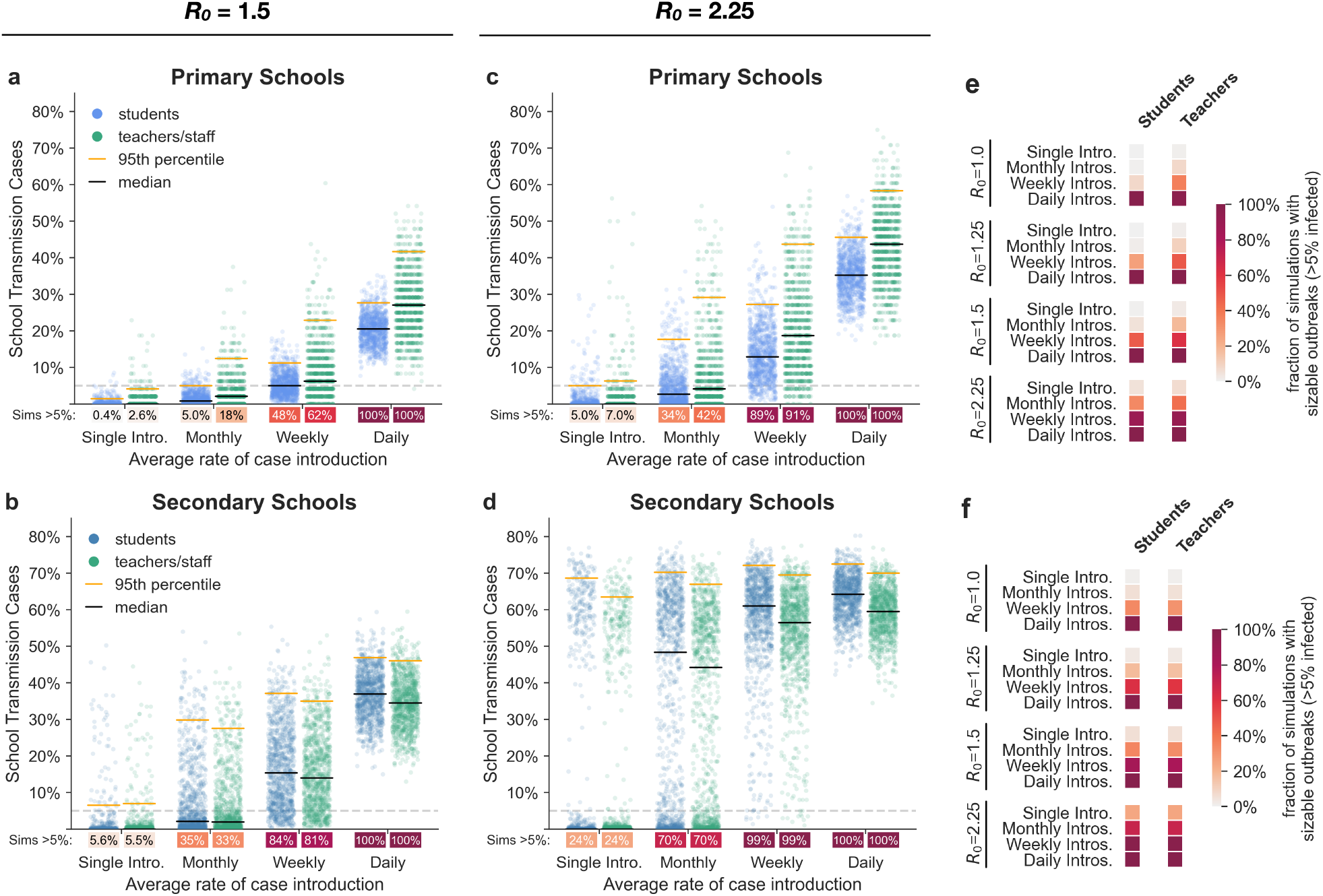
Effect of community prevalence. The distributions of school transmission cases as a percentage of the school population when new cases are introduced at different average rates. In these simulations, all students are in school five days a week and there is no proactive testing. (**a**,**b**) Outcomes for primary schools and secondary schools, respectively, with baseline transmission *R*_0_=1.5. Black and orange lines represent median and 95th percentile outcomes, respectively. Under each jitter distribution we list the percentage of simulations where more than 5% (grey dashed line) of the population are infected in school. (**c**,**d**) Outcomes for primary and secondary schools in scenarios with heightened transmission *R*_0_=2.25 due to the predominance of a highly-transmissible strain. (**e**,**f**) Heatmaps show the fraction of simulations where more than 5% of the student or teacher population are infected in primary and secondary schools, respectively, across a range of *R*_0_ values and introduction rates.

The probabilities of sizable outbreaks are higher in secondary schools than in primary schools, and the outbreaks tend to be larger in secondary schools. This difference holds across the range of parameters and interventions that we explore, and is primarily attributable to the difference in susceptibility between primary and secondary school students (Appendix B.3). Model outcomes are less sensitive to differences in the contact networks that characterize these settings (Appendix B.3.2).

### The effect of highly-transmissive variants

As of Spring 2021, several SARS-CoV-2 variants have evolved higher transmissibility relative to their ancestors.^35,42–47^ For example, the B.1.1.7 lineage that emerged from the United Kingdom appears to be 30%-70% more transmissible than previous SARS-CoV-2 variants.^35,42–44^ To understand how highly transmissible variants may impact transmission dynamics where they become predominant, we look at the consequences of a 50% increase in transmissibility, which increases the assumed baseline *R*_0_ for the school environment from *R*_0_=1.5 to *R*_0_=2.25. Results for more incremental increases in mean transmission rates that approximate intermediate penetrance of such strains can be found in Appendix B.1.1.

Figure 3 illustrates how community prevalence, as modeled by introduction rate, influences school transmissions when schools are confronted by this more transmissible strain. Even under a monthly rate of new case introductions, schools face the risk of a major outbreak. With more frequent introductions, substantive outbreaks become the most likely outcome. Aggressive controls mitigate the risk somewhat, but are considerably less effective for a strain with *R*_0_=2.25 than for a strain with *R*_0_=1.5 (Figure 5).

### The effects of interventions

#### Cohorting

Cohorting, wherein students are divided into two or more groups that alternate in-person learning, is a common strategy for mitigating outbreaks in school settings.^48–50^ In our model, we represent cohorting by shifting the contact networks according to which students are on campus (Appendix A.2.4.5). While off campus, students are disconnected from the school network but maintain household connections and transmission to casual contacts (the latter representing out-of-school interactions among the student body). Teachers remain on campus across all cohorts.

Figure 4 shows the effects of three common cohorting strategies: (1) all students belong to a single cohort that is on campus full time, (2) students are divided into two cohorts, A and B, which are on campus on alternating days, and (3) students are divided into two cohorts which are on campus on alternating weeks (Appendix A.2.6.3). We find that relative to no cohorting, both alternating day and alternating week strategies can improve outcomes substantially. Cohorting with alternating weeks generally outperforms cohorting with alternating days, but the marginal benefit of weekly cohorting is small when in-school transmission is limited (Appendix B.1.1). In primary schools, student cohorting alone dramatically reduces the risk of outbreak amongst students. In secondary schools, cohorting is helpful but insufficient on its own to keep the likelihood of an outbreak low amongst students or amongst teachers and staff.

**Figure 4:**
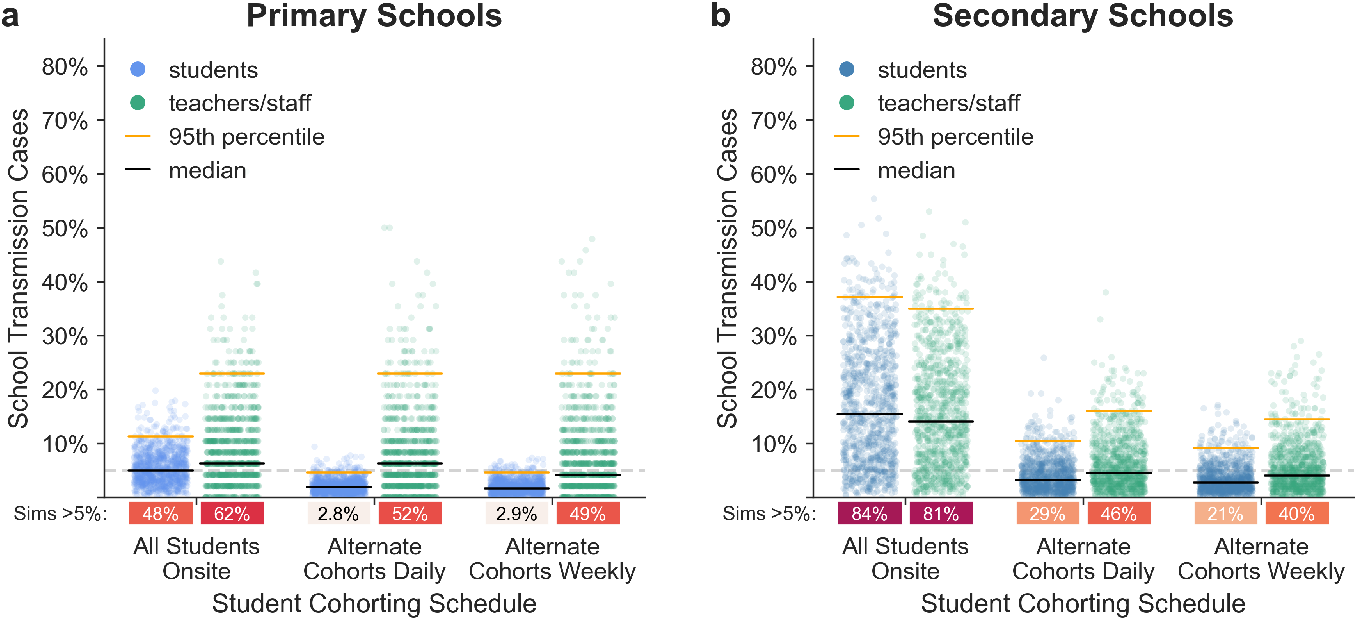
Effects of cohorting strategies. The distributions of school transmission cases as a percentage of school population for 1,000 simulations under different student cohorting strategies in (**a**) primary schools and (**b**) secondary schools with *R*_0_=1.5, approximately weekly new case introductions, and no testing. Under each jitter distribution we list the percentage of simulations that result in outbreaks affecting more than 5% of the population.

#### Proactive Testing

The purpose of proactive testing is to identify individuals who are infected but not currently showing symptoms, so that they can be quarantined.^51,52^ We consider five proactive testing strategies, detailed in Appendix A.2.6.2: (1) a baseline of no testing, (2) once-weekly testing amongst teachers and staff only, (3) twice-weekly testing amongst teachers and staff only, (4) once-weekly testing cadence amongst students, teachers, and staff, and (5) twice-weekly testing amongst students, teachers, and staff. We assume that 75% of students and 100% of teachers and staff are compliant with testing.^53^ Previous work suggests that long test turnaround times severely curtail the value of testing.^51,54,55^ To account for this, we use a test turnaround time of 24 hours and assume positive individuals enter quarantine immediately. In our basic model, only those who test positive are quarantined. We will later consider the consequences of quarantining all members of a primary school classroom when an individual therein tests positive.

Figure 5 illustrates the effects of cohorting and testing on the probability of sizable outbreaks. In our model, proactive testing consistently reduces the risk of outbreaks among teachers and students (Figure 5, Appendix Appendix B). While cohorting alone does not completely mitigate the risk of sizable outbreaks in secondary schools, the combination of cohorting and testing can keep this risk in check when baseline transmissibility in the school is sufficiently low.

**Figure 5:**
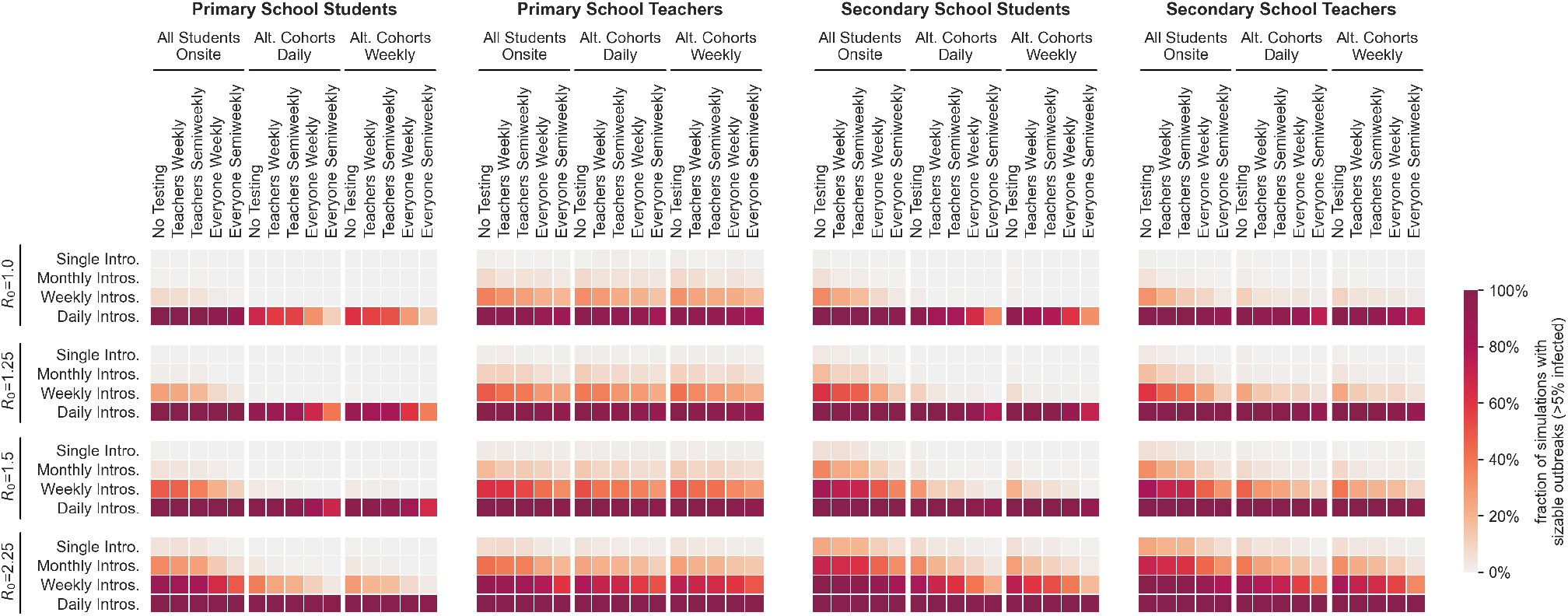
Effects of cohorting and testing strategies. Heatmaps illustrate the interactions of three student cohorting strategies and five proactive testing strategies (horizontal axis) across a range of transmission levels (*R*_0_) and new case introduction rates (vertical axis). The color of each cell indicates the fraction of 1,000 simulations for the given parameter set that result in sizable outbreaks where more than 5% of the population is infected. Outcomes are shown for student and teacher populations in primary and secondary schools as indicated by the title above each heatmap.

Figure 6 highlights the interactions between testing and cohorting measures in their effect on outbreak size in a secondary school environment. More aggressive testing helps reduce the size of outbreaks, as does cohorting. Testing and cohorting together outperform either measure alone. Interventions that help students also help teachers, and vice versa.

**Figure 6:**
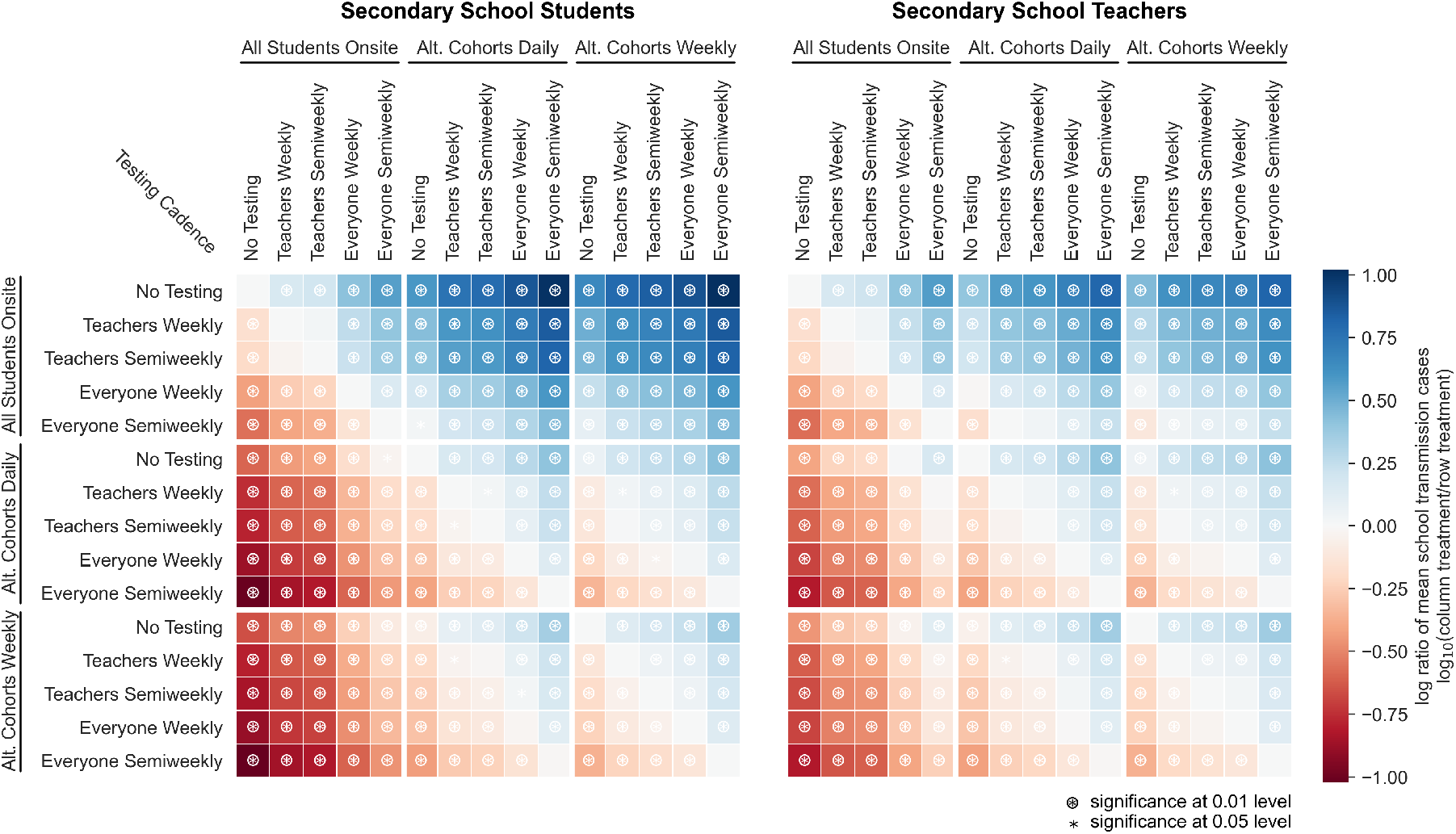
Relative effects of testing and cohorting in a secondary school setting. A heatmap of pairwise comparisons of testing and cohorting interventions illustrates the effects of various combinations on mean outbreak sizes. Each cell is colored according to the log-ratio of mean outbreak sizes for the two interventions, which represents the effect of the column intervention relative to the row intervention A **blue** cell indicates that the column intervention achieves a lower mean outbreak size than the row intervention, a **red** cell indicates that the column intervention has worse outcomes than the row intervention on average. Symbols in cells denote statistically significant differences in outbreak size distributions according to the Mann-Whitney U test at the 0.01 (⊛) and 0.05 (∗) levels. Results are shown for scenarios where *R*_0_=1.5, case introductions occur weekly on average, and only the positive individual is quarantined when cases are detected.

#### Isolation protocols

When an infected individual is identified by proactive testing, that person should be immediately isolated to prevent further transmission. In primary schools where classroom organization is stable, school administrators may additionally consider quarantining the entire classroom—students and teacher.

Our model indicates that classroom-level quarantine can reduce outbreak risk (Figure A4). For students and for teachers, with weekly introductions, the distribution of outcomes from isolating classrooms is stochastically smaller than the distribution of outcomes from isolating individuals (Mann-Whitney U test, *p* ≪ 0.01). This means that if one takes the outcome of a randomly drawn simulation run with classroom-level isolation and another with individual-level isolation, the classroom-level simulation run is significantly more likely to have the better outcome. When introductions are less frequent, benefits of classroom isolation may not be statistically significant. One important consideration for quarantining at the classroom-level is that this approach imposes more quarantine days on the population. When the level of in-school transmission is relatively low (e.g., *R*_0_ = 1.5), classroom-level isolation confers about the same amount of risk mitigation as moving from a weekly to semiweekly testing cadence, but classroom isolation leads to a large increase in the number of in-person learning days lost (Appendix B.9).

#### Vaccination

Pfizer-BioNTech and Moderna have reported extremely successful results from their phase III and phase IV COVID-19 vaccine trials, with 90% or greater efficacy at blocking symptomatic disease.^56–61^ The Johnson & Johnson vaccine has somewhat lower efficacy but requires only a single dose. Distribution of all three vaccines is well underway in the United States, with over a third of the US population fully vaccinated as of early May 2021.

Teachers and staff who have been vaccinated against COVID-19 are well-protected against infection (Figure 7). While initial phase III trial data focused only on diagnosis of symptomatic disease as a primary endpoint, current evidence suggests that the vaccines block transmission as well as symptomatic disease.^59,60,62,63^ Vaccinating teachers can also reduce the risk of outbreaks among students, particularly when paired with cohorting. The combination of vaccinating teachers and cohorting students continues to substantially reduce the risk of outbreaks at higher levels of transmissibility, which suggests this strategy may offer a proactive defense against the spread of more transmissive variants.

**Figure 7:**
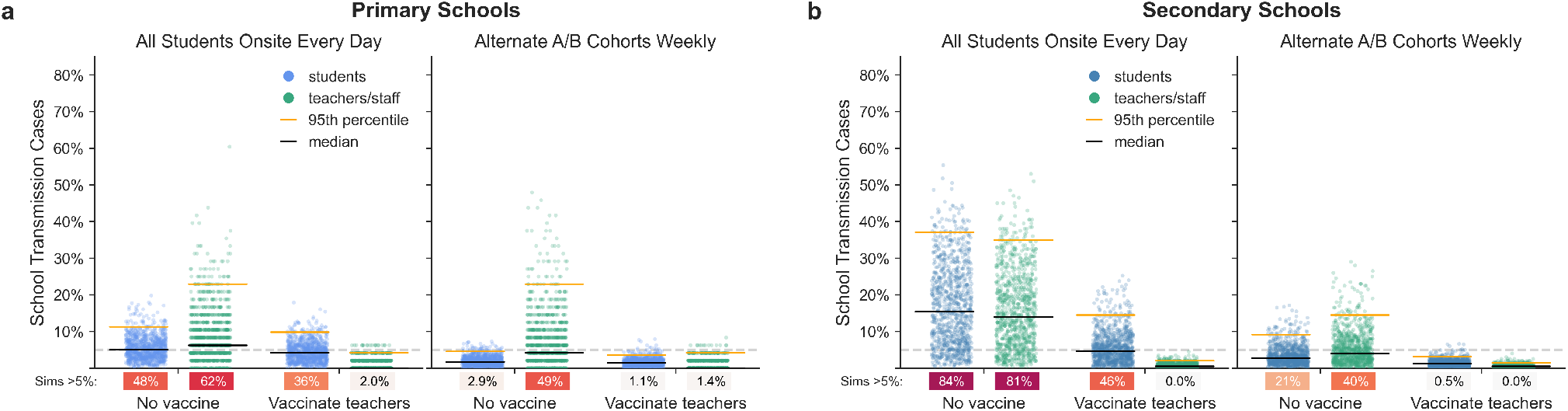
Effects of vaccinating teachers. The distributions of school transmission events as a percentage of school population for 1,000 simulations with either no vaccination or vaccination of all teachers and staff in (**a**) primary schools and (**b**) secondary schools. Results are shown for scenarios with *R*_0_=1.5, approximately weekly new case introductions, and no testing. Because vaccination is only 90% effective in the model, some teachers and staff become infected even when all are vaccinated. Effective vaccinations block both disease and transmission.

## Limitations

Like all epidemiological models, ours is a simplification of a complex, highly variable world. Our model is built on a series of assumptions and parameters; to the degree that these do not accurately reflect the real world, the model will be ineffective at predicting even the range of possible outcomes. We have attempted to account for uncertainty by embracing realistic heterogeneity and stochasticity in our model and by evaluating the sensitivity of outcomes across plausible ranges of values for critical parameters (Appendix Appendix B). Still, in a novel pandemic where many epidemiological parameters remain uncertain, and social and behavioral factors are fluid, some mismatch is inevitable.

The basic reproduction number (*R*_0_)—the average number of new cases generated by an infectious individual in a fully susceptible population—is a critical driver of disease dynamics. Estimates of *R*_0_ at the community level reflect an average rate of transmission integrated over many contexts and behaviors, which may include efforts to curtail transmission, such as social distancing, restricting large groups, and closing schools, businesses, and other gathering places. As schools reopen, larger numbers of individuals come together to interact, and average rates of transmission could be higher in schools than in the overall community. Still, basic in-school interventions such as mask wearing, physical distancing, and behavioral changes are expected to substantially reduce *R*_0_.

In our model, we assume that these basic interventions can reduce *R*_0_ to 1.5—roughly half of what it would be in the absence of intervention, depending on the transmissibility of variants circulating in the population. Previous studies suggest that transmission is relatively limited in schools.^22,49,50,64–66^ In addition to the basic measures listed above, many of the schools described in these studies were already implementing one or more interventions along the lines of the ones we analyze here: cohorting, isolating groups, testing, contact tracing, reducing the number of people on campus, and so forth.^17,49,66,67^ These studies largely corroborate our findings that school transmission is often kept in check when such mitigation strategies are used. Fewer studies have considered schools that are only using masks and other basic measures, but there is evidence that sizable school outbreaks can occur in these contexts.^18^ We find that the probability and size of outbreaks are strongly influenced by the underlying *R*_0_, but the relative effects of mitigations are robust across a range of *R*_0_ values (Appendix B.1.1).

Here we have simulated a subset of the currently practiced strategies for returning to in-person learning.^68^ We assume that students, teachers and school staff adhere to testing cadences, cohorting schedules, and quarantine policies in addition to basic measures. In the absence of evidence to the contrary, we assume that—holding transmissibility and susceptibility constant—all forms of close contact are equally likely to result in transmission. In practice, the nature of interpersonal relations may make transmission from student to student or from teacher to teacher more likely than transmission between these groups, and could explain why some contact-tracing studies have reported disproportionately low student-to-teacher transmission.^49,69^

We have modeled primary school children as being less susceptible to SARS-CoV-2 infection than teachers and staff. Recent evidence from seroprevalence and contact-tracing studies support this assumption.^16,20,70–72^ However, because a high percentage of children develop asymptomatic disease, COVID-19 cases among children may be more likely to go undetected. Therefore, it is possible that the apparent decreased susceptibility to SARS-CoV-2 infection among primary school-age children is an artifact of underreporting. We find that school transmission is sensitive to the susceptibility of students, and if the susceptibility of primary school students is closer to that of adults then primary school outcomes will more closely resemble those of secondary schools (Appendix B.3.1).

We also assume that there is no difference in infectiousness between pre-symptomatic, asymptomatic, and symptomatic individuals. Some studies suggest that pre-symptomatic individuals may contribute a disproportionately high number of cases relative to the duration of this disease state, and asymptomatic individuals may contribute disproportionately few cases relative to symptomatic individuals.^73–75^ However, these studies draw on contact tracing data, and it is unclear how much of these differences in transmission are attributable to differences in viral transmissibility (e.g., viral load and shedding) as opposed to changes in behavior or other factors associated with these disease states. Our assumption is conservative with respect to asymptomatic transmission in the sense that our results will err on the side of overestimating the number of school transmission events if asymptomatic or pre-symptomatic individuals are indeed less infectious than symptomatic individuals. However, our results may overstate the relative benefit of mitigations in such a case. If pre-symptomatic individuals are more infectious than symptomatic individuals, our results may underestimate the amount of transmission that is likely to occur in schools, but the importance of proactive mitigations would be even greater. The sensitivity of our model results to the relative infectiousness of pre-symptomatic and asymptomatic individuals is discussed in Appendix B.1.3.

Over the course of the pandemic, community prevalence measurements have fluctuated substantially on a timescale of months, due to changing individual behaviors and societal interventions. Because these fluctuations have been largely unpredictable, we have elected to use a constant introduction rate throughout. In doing so we are effectively decoupling infection dynamics within the school from epidemic dynamics in the community. In our model, intervention choices that lead to a large number of school-related transmissions do not feed back on the community prevalence to influence the downstream hazard of community introduction back into the school. Similarly, in our model, mitigation choices that block school transmission do not reduce the community introduction rate. This seems reasonable when schools are not important drivers of the community prevalence of SARS-CoV-2 infection, as appears to be the case especially for K-5 schools.^17,49,76^ Where schools are important drivers of community dynamics, however, our model risks underestimating the consequences of mitigation efforts. When schools drive community prevalence, planners must also consider the cost of the additional community infections that result from reopening schools—which we have not done here.

## Summary

We have presented results from a simulation model of reopening schools during the COVID-19 pandemic. The purpose of this model is to provide a scenario-simulating tool that, when used in concert along with other credible sources of information and data, can aid decisions around school reopening policies.

We attempt to make reasonable assumptions about epidemiological parameters and aspects of human behavior that drive disease transmission. Our results tend to be robust to these choices, and the qualitative findings that we report—advantages to cohorting, testing, and vaccination—are expected to hold up more broadly. In Appendix B, we provide detailed sensitivity analyses for a number of important parameters, including transmissibility, student susceptibilities, contact network structures, and compliance with intervention strategies. Our online webapp (https://www.color.com/impact-of-primary-school-covid-19-testing) provides a way to explore the range of parameters interactively, which can be used to assist in dynamic decision-making in response to uncertain and changing local circumstances.

Our model suggests that dividing students into cohorts that attend school in person on alternating schedules can be a powerful strategy for mitigating risk. Cohorting is effective in our model because students largely restrict in-person interactions to other individuals within their own groups, and this takes place only while at school. The cohorting strategy is fairly robust to students interacting off campus as well, provided that students continue to limit their contacts to students in their own cohort (Appendix B.7). However, when students socialize beyond their close contacts and across cohort boundaries outside of school—as students are wont to do—the effectiveness of cohorting is reduced (Appendix B.2). Schools could consider further efforts to reduce the mixing of the student body at school, which has a significant impact on the risk of transmission in all contexts (Appendix B.2). This might include restructuring lunch periods, passing periods, transportation logistics, and other scenarios in which incidental transmission could occur between otherwise “unconnected” individuals.

Teachers and staff are more susceptible to the virus than primary school students and at higher risk of severe disease than students of any age. Moreover, teachers serve as conduits for outbreaks to move among classrooms within the school network. Frequent, proactive testing of teachers and staff can interrupt such transmission chains and further protect them from infection.

Vaccinating teachers and staff is a powerful tool for protecting this critical workforce. If vaccines effectively block SARS-CoV-2 transmission in addition to COVID-19 symptoms, vaccinating teachers and staff can significantly dampen outbreak dynamics in both primary and secondary schools. The result would be fewer cases among adults and students alike.

The success of reopening efforts will hinge on the amount of transmission that occurs in schools. The higher the transmissibility, parameterized here as *R*_0_, the greater the chance of substantial outbreaks in a school setting. Physical distancing, diligent use of masks, and other environmental controls offer a first-line approach to reducing transmission and will be an important component of reopening plans.

For both primary and secondary schools, the risk of an outbreak increases as cases in the surrounding community rise. One of the most effective ways to safely reopen schools is by controlling COVID-19 in the community. Surveillance should be in place to monitor levels of community transmission and schools should be prepared to respond flexibly.

Because highly transmissible variants such as B.1.1.7 pose increased risks for outbreaks, schools need to be vigilant on multiple fronts. First, where genomic surveillance is available, school districts and counties need to monitor the introduction and spread of these variants. Second, irrespective of the variants involved, it will be important to monitor epidemic dynamics within any given school and to respond quickly should uncontrolled spread take place. An additional virtue of testing is that it facilitates early detection of such events. We have not explicitly modeled surveillance testing and response, but general public health guidance should be followed. For example, schools could implement “tripwire” strategies, returning to distance learning for a period of time in response in-school outbreaks or rising community prevalence. In the event of isolated cases appearing at higher than expected rates, administrators should reconsider assumptions about the rate of community introduction and intensify control measures accordingly.

Our model suggests that under certain parameters, it may become difficult or impossible to keep the probability of outbreaks low across the schools of an entire district. Tripwire strategies may be necessary under these circumstances.

While gaps remain in our understanding of transmission in school settings, both real-world experience and models — including the one presented here — suggest a path forward for schools to reopen, particularly when community transmission is low and when it is possible to deploy and consistently implement the mitigation measures we have modeled here.

## Data Availability

Extensive documentation and code for the SEIRS+ framework that was developed and used to implement the Extended SEIR Network Model studied in this work can be found at github.com/ryansmcgee/seirsplus. The datasets generated by this work can be explored using an interactive web applet hosted at color.com/impact-of-primary-school-covid-19-testing. Simulation and statistics data will be accessible via the aforementioned github and otherwise made available upon request.

https://www.color.com/impact-of-primary-school-covid-19-testing

## Acknowledgements

The authors thank Martin Rosvall for help in developing the contact network structures used in the SEIRS+ model. Ted Bergstrom, Natalie Dean, Bill Hanage, Michael Lachmann, and Marc Lipsitch provided valuable feedback in developing the model and adapting it to the school scenarios considered here.

## Author contributions

Conceived of the model: RSM, CTB. Reviewed the literature: RSM, HEW. Parameterized the model: RSM, JRH, HEW. Implemented the model and ran the simulations: RSM. Produced the data visualizations: RSM, JRH. Analyzed the results: RSM, CTB, JRH, HEW, AYZ. Developed the interactive web app: JRH. Drafted the manuscript: CTB, RSM, HEW, JRH, AYZ.

## Disclosures

CTB and RSM consult for Color Health. CTB has received honoraria from Novartis. JRH, HEW and AYZ are currently employed by and have equity interest in Color Health.

## Notes

### Funding Statement

CTB and RSM were paid consulting fees by Color Health.

### Summary of Updates

Text edited for clarity in some places; presenting a thorough sensitivity analysis with results mentioned throughout the main text and full details presented in a new Appendix B.

