## Appendices for "Model-driven mitigation measures for reopening schools during the COVID-19 pandemic"

### Appendix A Model Description

#### A.1 SEIRS+ Extended SEIR Network Model

SEIRS+ is an open source Python framework developed by McGee et al. that supports flexible parameterization and implementation of sophisticated epidemiological models (https://github.com/ryansmcgee/seirsplus). The models studied in this work are parameterizations of the stochastic Extended SEIR Network Model provided in the SEIRS+ framework. We simulate our models using the Interventions Simulation Loop provided in SEIRS+ with minor modifications for our particular school context. Extensive documentation for the models, simulation loops, and other features of SEIRS+ can be found on the SEIRS+ github wiki (https://github.com/ryansmcgee/seirsplus/wiki).

##### A.1.1 Heterogeneity

In the SEIRS+ Extended SEIR Network Model, individuals are represented as nodes in a contact network, and all parameters, interactions, and interventions can be specified on a node-by-node basis. Therefore, this model enables explicit representation of heterogeneity in disease characteristics, contact patterns, and behaviors, which are important for modeling small, age-stratified populations such as schools. Parameter choices and distributions for our school models are described in the School Models appendix section.

##### A.1.2 Compartments

The Extended SEIR Network Model extends the classic SEIR model of infectious disease to represent pre-symptomatic, asymptomatic, and symptomatic disease states, which are of particular relevance to the SARS-CoV-2 pandemic. The classic SEIR model divides the population into susceptible ($S$), exposed ($E$), infectious ($I$), and recovered ($R$) individuals. In this extended model, the infectious subpopulation is further subdivided into pre-symptomatic ($I_{\text{pre}}$), asymptomatic ($I_{\text{asym}}$), and symptomatic ($I_{\text{sym}}$) compartments, all of which represent contagious individuals (the full Extended SEIR Network Model includes a hospitalized infectious state, but we assume no hospitalization in this report and effectively ignore this compartment). Individuals transition from one compartment to the next at times determined by the disease characteristics (see Appendix A.2.2 Disease progression parameters). A parameterizable fraction of infected individuals are deemed asymptomatic and will progress to the asymptomatic compartment when exiting the presymptomatic compartment, while the remainder of infected individuals will progress to the symptomatic compartment. The dynamics of compartment transitions are described further in the Dynamics appendix section.

The effect of isolating individuals in response to symptoms or testing is modeled by introducing compartments that represent quarantined individuals (Figure A1). An individual may be quarantined in any disease state, and every disease state has a corresponding quarantine compartment. Quarantined individuals follow the same progression through the disease states, but their set of close contacts are defined by a distinct quarantine contact network (Appendix A.2.4 Contact Networks). In this work, individuals are moved into quarantine states by the Intervention Simulation Loop (Appendix A.2.6.1), such as when a positive test result is returned, as opposed to according to a transition rate. Individuals remain in the quarantine compartment flow until the designated isolation

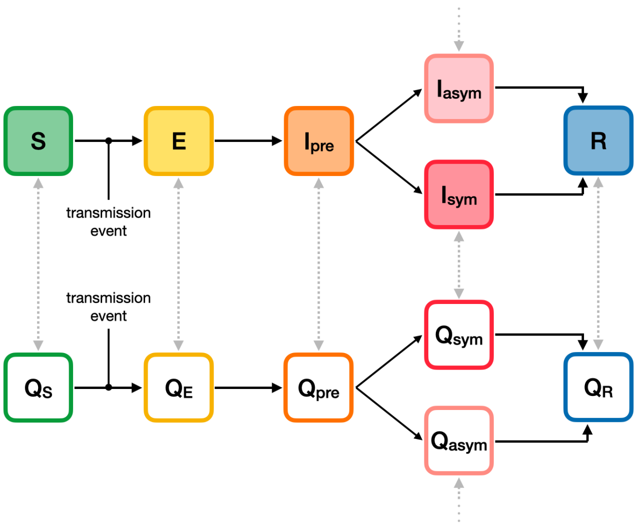

**Figure A1: Compartment model.** The compartment model that defines the progression of disease states in the Extended SEIR Network Model.

period has been reached (10 days in this work), at which time they are moved into the non-quarantine compartment corresponding to their current disease state.

##### A.1.3 Dynamics

Transmission dynamics are simulated using the Gillespie algorithm, a common and rigorous method for simulating stochastic interaction dynamics. Briefly, the system's differential equations are adapted to compute the 'propensity' of the possible events (i.e., the expected amount of time until a given event will take place) for all nodes at each time step. These propensities are then used to compute the probabilities of all possible state events normalized across the entire population. A random node and corresponding transition are selected to execute according to these probabilities in each time step. The propensities of transmission events (S to E transitions) are proportional to the product of the prevalence of infectious individuals among each node's contacts and the transmissibilities and susceptibilities of the interacting individuals (see Appendix A.1.4 Transmission).

SEIRS+ supports calculating the propensities of disease progression transitions (e.g., $E$ to $I_{\text{pre}}$, $Q_{\text{pre}}$ to $Q_{\text{sym}}$) in two different ways: 1) using transition rates (standard Gillespie implementation), or 2) using compartment residence times. Running the model in the first mode results in exponentially distributed residence times in each compartment, as in classic mass action SEIR-like models. In the latter mode, each individual is assigned a residence time for each compartment, and the propensity for a given individual to transition out of the current state is 0 until this residence period has elapsed, at which time this propensity becomes large such that the next event will be this given individual transitioning to the next compartment with probability approaching 1. This results in a hybrid model where transmission events occur stochastically according to the Gillespie algorithm while other disease progression transitions occur in a clock-like manner in parallel. In reality, residence times in each disease state are not exponentially distributed, and the discrepancies can be particularly important when looking at early stages of an outbreak and when considering control strategies such as proactive testing.[77,78] As such, we use the residence time propensity calculation mode in this work and assign heterogeneous residence times to individuals drawn from gamma distributions that better match empirical descriptions of the disease dynamics for COVID-19 (Appendix A.2.2).

For more information about the propensity equations, refer to https://github.com/ryansmcgee/seirsplus/wiki.

##### A.1.4 Transmission

The dynamics governing transmission events that cause susceptible individuals to become exposed are fundamental to any epidemiological model, so we break down the transmission dynamics of the Extended SEIR Network Model in detail here.

In general, the propensity $P^{(i)}(S \to E)$ of a given susceptible individual $i$ becoming infected is proportional to the product of the prevalence of infectious individuals among their contacts, the average transmissibility of their infectious contacts $\overline{\beta}^{\text{(contacts)}}$, and their own susceptibility to infection $\alpha^{(i)}$.

$$P^{(i)}(S \to E) \propto \alpha^{(i)} \times \overline{\beta}^{\text{(contacts)}} \times \text{(prevalence among contacts)}$$

An individual's transmissibility $\beta^{(i)}$ (i.e., transmission rate) is equal to the expected number of cases that this individual would generate in a fully-susceptible population (i.e., the reproduction number for the individual, $R_0^{(i)}$

divided by the length of their infectious period $\gamma^{(i)}$.

$$\beta^{(i)} = \frac{R_0^{(i)}}{\gamma^{(i)}}$$

For the purposes of the models considered in this work, the propensity of a given individual $i$ becoming infected is calculated using the following equation[1], which we will break down in the rest of this section

$$P^{(i)}(S \to E) = \alpha^{(i)} \left[ p \underbrace{\left( \frac{\bar{\beta}\left(I_{\text{pre}} + I_{\text{sym}} + I_{\text{asym}}\right)}{N} \right)}_{\text{global transmission}} + (1-p) \underbrace{\left( \frac{\sum_{j \in C_G^{(i)}} \delta^{(ji)} \left( \beta^{(j)} \mathbf{1}_{X^{(j)} \in \{I_{\text{pre}}, I_{\text{sym}}, I_{\text{asym}}\}} \right)}{|C_G^{(i)}|} \right)}_{\text{local transmission}} \right]$$

In this model, disease transmission may occur either from close contacts defined by the contact network structure or from casual contacts. Close contacts are individuals with whom one has repeated, sustained, or close proximity interactions on a regular basis: classmates, friends, housemates, or other close relationships. In contrast, casual contacts are individuals with whom one has incidental, brief, or superficial contact on an infrequent basis and to whom one is not connected directly on the network. A network locality parameter $p$ sets the relative frequency and weight of transmission among close (local network) and casual (global) contacts in the model population.

**A.1.4.1 Global transmission** A fraction $p$ of a given individual's interactions are with casual contacts, which are assumed to be individuals randomly sampled from the population at large, irrespective of the contact network. With respect to these global interactions, every node in the population is equally likely to come into contact with every other node, and the population can be considered well-mixed. Thus the propensity of global transmission is calculated in the same way as mass action compartment models that assume a well-mixed population. The propensity for a given susceptible individual to become exposed due to global transmission is proportional to the product of that individual's susceptibility $\alpha^{(i)}$, the population mean transmissibility of infectious individuals $\overline{\beta}$, and the prevalence of infectious individuals in the overall population $(I_{\text{pre}} + I_{\text{sym}} + I_{\text{asym}})/N$.

**A.1.4.2 Local transmission** A fraction $1-p$ of a given individual's interactions are with individuals from their set of "close contacts." An individual's close contacts are defined as the nodes adjacent to the given node in the contact network. $C_G^{(i)}$ denotes the set of close contacts for individual $i$: the nodes adjacent to node $i$ in the contact network graph $G$. $|C_G^{(i)}|$ denotes the size of this set: the number of close contacts that $i$ has.

With respect to local transmission, transmissibility is considered on a pairwise basis. That is, every directed edge of the contact network representing transmission from infected node $j$ to susceptible node $i$ is assigned a transmissibility weight $\beta^{(j)}$ that depends on the transmissibility of the infected individual $j$ alone. The propensity for a given susceptible individual to become exposed due to local transmission is calculated as the product of that individual's susceptibility and the sum transmissibility of their infectious close contacts ($\mathbf{1}_{X^{(j)} \in \{I_{\text{pre}}, I_{\text{sym}}, I_{\text{asym}}\}}$ is an

[1]This equation is simplified from the general equation implemented in the Extended SEIR Network Model, which includes parameters and terms that are not used here and are thus zeroed out.

indicator function that takes the value 1 when the state $X^{(j)}$ of the contact node $j$ is one of the infectious states and 0 otherwise), scaled by the size of their local network as described below ($|C_G^{(i)}|$ denotes the size of the set of close contacts for individual $i$).

This amounts to the propensity of exposure for node $i$ being proportional to the product of their susceptibility and the transmissibility-weighted prevalence of infectious individuals in their local network. Thus, propensity for exposure due to local transmission is frequency dependent and analogous to the propensity contribution from global transmission. Implicit in this formulation is an assumption that all individuals have limited interaction budgets and individuals with more close contacts (i.e., higher degree) interact less with each contact and are therefore less likely to become exposed by any single individual. A factor $\delta^{(ji)}$ appears in the calculation of propensity for exposure due to local transmission. This pairwise factor is used to re-weight the transmissibility of interactions according to the connectivity of the interacting individuals. While it is reasonable to think that individuals (e.g., secondary school teachers) who have many contacts (e.g., students) do not interact as closely with each of their contacts as another individual who only has a handful of contacts, we do not assume that the propensity of infection decreases linearly with degree for SARS-CoV-2 transmission. We define the degree scaling factor $\delta^{(ji)}$ as

$$\delta^{(ji)} = \frac{\log(D^{(i)}) + \log(D^{(j)})}{2\log(\overline{D})},$$

where $D^{(j)}$ and $D^{(i)}$ are the degrees of nodes $j$ and $i$, respectively, and $\overline{D}$ is the mean degree of the network. Thus, the propensity for infection by a single infectious contact is lower for highly-connected individuals compared to low connectivity individuals, but not proportionally so.

#### A.2 School Models

The following sections describe the specific assumptions and parameter values used to define the primary and secondary school models studied in this work. These models were implemented using the SEIRS+ framework's Extended SEIR Network Model (see Appendix A.1 SEIRS+ Extended SEIR Network Model).

##### A.2.1 Parameters Overview

Table A.2.1 gives an overview of parameter values used in the primary and secondary school models. More information about these parameters can be found throughout Appendix A. Exploration of the sensitivity of the models to these parameters can be found in Appendix B.

**Table A.2.1** Overview of parameter values for the primary and secondary school models.

| **Parameter** | **Primary School Model** | **Secondary School Model** |
|---|---|---|
| Basic reproduction number $R_0$ | 1.5 or 2.25 (highly transmissible variant)<br>*See Appendix B.1.1* | 1.5 or 2.25 (highly transmissible variant)<br>*See Appendix B.1.1* |
| Individual $R_0^{(i)}$ coefficient of variation | 2.0 (overdispersed)[27–29]<br>*See Table A.2.3*<br>*See Appendix B.2* | 2.0 (overdispersed)[27–29]<br>*See Table A.2.3*<br>*See Appendix B.2* |
| Presymptomatic transmissibility<br>*(relative to symptomatic transmissibility)* | 100%<br>*See Appendix B.1.3* | 100%<br>*See Appendix B.1.3* |
| Asymptomatic transmissibility<br>*(relative to symptomatic transmissibility)* | 100%<br>*See Appendix B.1.3* | 100%<br>*See Appendix B.1.3* |
| Susceptibility of students<br>*(relative to susceptibility of adults)* | 60%[16,20]<br>*See Appendix B.3.1* | 100%[16,20]<br>*See Appendix B.3.1* |
| Fraction of asymptomatic cases | 40% of students,[22,37,79]<br>30% of teachers/staff[80,81] | 30% of students,[80,81]<br>30% of teachers/staff[80,81] |
| Disease state periods | *See Table A.2.2* | *See Table A.2.2* |
| Compliance with isolation upon symptoms | 20%<br>*See Appendix B.4* | 20%<br>*See Appendix B.4* |
| Compliance with testing | 75% of students,[53]<br>100% of teachers/staff | 75% of students,[53]<br>100% of teachers/staff |
| Test sensitivity | RNA-based time series[82]<br>*See Appendix A.2.6.2* | RNA-based time series[82]<br>*See Appendix A.2.6.2* |
| Vaccine uptake | 0% of students,<br>0 or 100% of teachers/staff | 0% of students,<br>0 or 100% of teachers/staff |
| Vaccine effectiveness | 90%[56–61]<br>*See Appendix B.8* | 90%[56,57,59–61]<br>*See Appendix B.8* |
| Rate of disease with effective vaccine | 0%[56–61] | 0%[56,57,59–61] |
| Rate of transmission with effective vaccine | 0%[62,63] | 0%[62,63] |
| Contact network parameters | *See Table A.2.4a, Table A.2.4b* | *See Table A.2.4c, Table A.2.4d* |
| Proportion of global transmission | 20%[26]<br>*See Appendix B.2* | 20%[26]<br>*See Appendix B.2* |

##### A.2.2 Disease progression parameters

As described in Appendix A.1.3 Dynamics, individuals remain in each compartment (excluding Susceptible) for a designated period of time before progressing to the next disease state. The population is heterogeneous for each disease state period, with each individual being assigned disease state periods drawn from gamma distributions that are informed by empirical studies of COVID-19 progression. Refer to Table A.2.2 for more information about each distribution. We assume that the distributions of disease state periods are the same for all age groups and for both quarantined and non-quarantined individuals. The same gamma distribution parameters are used to define the period probability distributions in every simulation, but the period values are randomly drawn and assigned in each replicate.

Additionally, we assume that 30% of adults and secondary school students are asymptomatic, and that 40% of primary school students (young children) are asymptomatic. In the initialization of each simulation, each individual in the population is randomly assigned a symptomatic or asymptomatic status according to these probabilities. If an individual becomes infected, they will progress to the symptomatic ($I_{\text{sym}}$) or asymptomatic ($I_{\text{asym}}$) state when exiting the pre-symptomatic ($I_{\text{pre}}$) state according to this assigned status. 20% of symptomatic individuals self-isolate upon entering the symptomatic state, but there are no other parameter differences between symptomatic and asymptomatic individuals in our model.

**Table A.2.2** A representative distribution of period values drawn for a secondary school with 1,000 individuals is shown for each parameter in the center column below. Statistics across all replicate distributions in our analysis are shown in the rightmost column.

<table>
<tr><th>Disease state period</th><th>Distribution</th><th>Statistics</th></tr>
<tr><td>Latent period<br>(time in $E$ state)</td><td>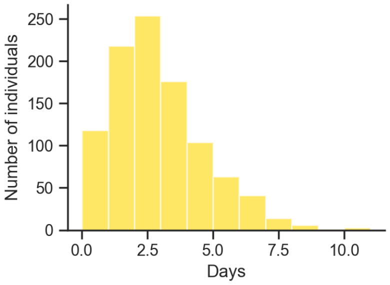

gamma(mean=3.0, CV=0.6)</td><td>mean 3.0 days<br>std 1.8 days<br>95% CI (0.6, 7.4)<br><br>References: [30,83–87]</td></tr>
<tr><td>Pre-symptomatic period<br>(time in $I_{pre}$ state)</td><td>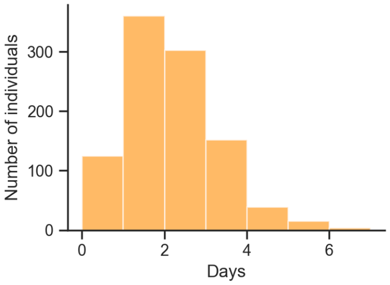

gamma(mean=2.2, CV=0.5)</td><td>mean 2.2 days<br>std 1.1 days<br>95% CI (0.6, 4.8)<br><br>References: [83,84]</td></tr>
<tr><td>Symptomatic period<br>(time in $I_{sym}$ or $I_{asym}$ state)</td><td>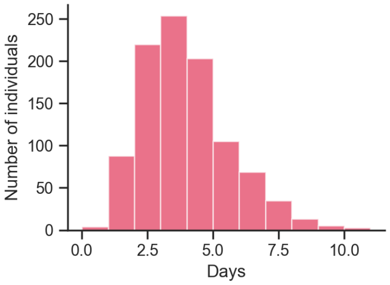

gamma(mean=4.0, CV=0.4)</td><td>mean 4.0 days<br>std 1.6 days<br>95% CI (1.5, 7.6)<br><br>References: [84,88–90]</td></tr>
<tr><td>Total infectious period<br>(total time in $I_{pre}$, $I_{sym}$, and $I_{asym}$ states)</td><td>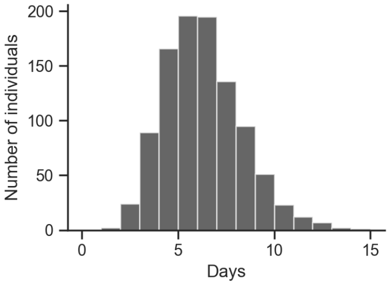

gamma(mean=2.2, CV=0.5)</td><td>mean 6.2 days<br>std 1.9 days<br>95% CI (3.0, 10.5)<br><br>References: [83,84,88–90]</td></tr>
</table>

##### A.2.3 Transmission parameters

As described in Appendix A.1.4 Transmission, the propensities of transmission events depend on the transmissibility and susceptibility parameters of interacting individuals. Each individual is assigned an individual reproduction number $R_0^{(i)}$, which is the expected number of secondary cases that the individual generates when infectious in a fully susceptible population. Each individual reproduction number is converted to an individual transmissibility (i.e., transmission rate) parameter using the following standard formula

$$\beta^{(i)} = \frac{R_0^{(i)}}{\gamma^{(i)}},$$

where $\gamma^{(i)}$ is the total infectious period for individual $i$. We assume that individual transmissibility is heterogeneous and follows an overdispersed (long-tailed) distribution that corresponds approximately to 20% of individuals contributing 80% of the total expected number of secondary cases (the "80/20 rule").[27–29] We calibrate the individual reproduction number distribution such that its mean corresponds to a chosen average basic reproduction number $R_0$ for the population ($R_0$=1.5 and $R_0$=2.25 are considered in the main text) and so that 80% of the weight falls in the upper 20th percentile of individuals in the tail of the distribution. Therefore, for any $R_0$ considered in this paper, many individuals are expected to generate fewer than 1 secondary case while a minority of individuals are expected to contribute a large number. Refer to Table A.2.3 for more information about these distributions. We assume that all age groups have transmissibilities drawn from the same distribution. In addition, we assume there is no difference in transmissibility between the pre-symptomatic, symptomatic, and asymptomatic states (i.e., the same individual transmissibility is used while an individual is in any one of these states).

Additionally, individuals are assigned a susceptibility parameter value, which weights the propensity that they become infected by any infectious contacts they may have (see Appendix A.1.4 Transmission). Adults and secondary students are assigned the baseline susceptibility value of 1.0, and thus their propensity of infection is based on the unweighted transmissibilities of their contacts. In the main text, primary school students (young children) are assumed to be 60% as susceptible as adults. Therefore, primary school students are assigned a susceptibility value of 0.6, and their propensity of infection is only 60% of that of an adult in the same infectious contact context.

Global transmission can be thought to represent both casual interactions among members of the school population while on campus as well as relatively infrequent interactions among members of the school population while off campus (e.g., on weekends and off-cohort days). We assume that 80% of transmission is attributable to transmission between close contacts and 20% is attributable to global transmission among casual contacts (see Appendix A.1.4 Transmission). This parameterization is supported by the extensive contact survey conducted by Mossong et al. (2008), which finds that approximately 20-30% of interactions where transmission is possible are with first-time contacts or last less than 15 minutes.[26]

**Table A.2.3** A representative distribution of drawn individual reproduction number values for a secondary school with 1,000 individuals is shown for the basic reproduction numbers considered in the main text below. Statistics across all replicate distributions in our analysis are shown in the rightmost column.

| **Population $R_0$** | **Distribution of individual reproduction numbers $R_0^{(i)}$** | **Statistics** |
|---|---|---|
| $R_0$ = 1.5 | 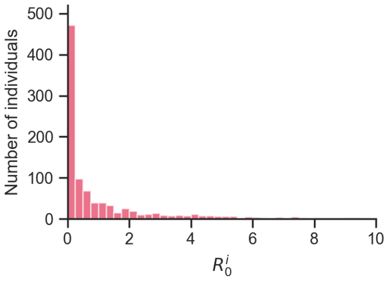<br><br>`gamma(mean=1.5, CV=2.0)` | 80% of weight in top 20th percentile.[27–29]<br><br>mean 1.5<br>std 3.0<br>median 0.26<br>95% CI (0, 10.2)<br>80th percentile: 2.2<br>31% of values > 1.0 |
| $R_0$ = 2.25 | 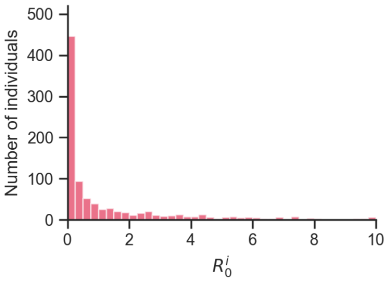<br><br>`gamma(mean=2.25, CV=2.0)` | 80% of weight in top 20th percentile.[27–29]<br><br>mean 2.25<br>std 4.5<br>median 0.39<br>95% CI (0, 15.45)<br>80th percentile: 3.3<br>38% of values > 1.0 |

##### A.2.4 Contact Networks

The SEIRS+ Extended SEIR Network Model allows arbitrary graphs to be used to specify the contact network that defines close contacts for local transmission (see Appendix A.1.4 Transmission). Here we define distinct networks representing the contact structure of a primary school and a secondary school.

**A.2.4.1 Primary school contact networks** For our primary school model, we simulate a medium-sized school of 480 students with 24 teachers and 24 additional staff. Each class comprises one teacher and 20 students in mutual contact. That is, the students and teacher for each classroom are strongly connected. Additionally, each teacher interacts with a handful of other teachers and staff, and students that share the same household are connected. The percentage of primary school aged children that share a household with another primary school aged child is calibrated by US census data. Most of the contacts that an individual makes in the school population are with the students and teacher in their own class, and disease transmission within a class is more likely than between classes. The FARZ algorithm generates random networks with built-in community structure and broad, heavy-tailed degree distributions that are realistic for human contact networks.[24,25,41,91,92] Refer to Table A.2.4a for more information about the parameterization of these networks, and see Table A.2.4b for more information about their degree distributions and other network properties.

**A.2.4.2 Secondary school contact networks** For our secondary school model, we consider a medium-sized school with 800 students (200 per graduating class), 125 teachers, and 75 staff. We generate network layers for students and teachers and staff using the FARZ network generation algorithm, which allows us to calibrate epidemiologically-important network properties (e.g., cluster structure, assortativity, and clustering coefficient) to values consistent with studies of secondary school contact networks.[38,39] A FARZ network layer is generated for each grade, with students belonging to one or more social groups (i.e., network clusters) of about 10 individuals each. 80% of each student's contacts are with students in the same grade, and 80% of those within-grade contacts are with students in their own social groups. Students that share a household are connected as well. The percentage of secondary school aged children that share a household with another secondary school aged child is calibrated by US census data. Interactions between teachers and staff are represented by another FARZ network layer. Finally, students are connected with six random teachers with whom they have classes. Each teacher is associated with a grade level, and students take classes with teachers in their own grade level 75% of the time, which leads to students in the same grade being more likely to share teachers. A unique random network is generated as described for each simulation replicate. Refer to Table A.2.4c for more information about the parameterization of these networks, and see Table A.2.4d for more information about their degree distributions and other network properties.

**A.2.4.3 Quarantine contact networks** When individuals are in quarantine, a separate quarantine contact network is referenced when calculating propensities of transmission involving that individual. Here we define the contact network as the school contact network with all edges removed except for those between housemates. That is, a quarantined individual makes contact with their housemates (e.g., siblings) but no one else from the school population. Global transmission is set to 0 for individuals in quarantine.

**A.2.4.4 Weekend contact networks** The contact network that is in effect on weekends is the same as the quarantine network. That is, individuals only have direct contacts with housemates on weekends. However, global transmission is left at 20% for non-quarantined individuals on weekends to represent general mixing among the school population when out of school.

**A.2.4.5 Cohort contact networks** One of the mitigations we consider is student cohorting, in which students are divided into two groups, only one of which attends school on any given day (see Appendix A.2.6.3 Cohorting). In our model, cohorting is implemented by alternating between two modified school contact networks. Students are divided into two cohorts, A and B. Primary students are divided such that exactly half of each classroom is in each cohort. Secondary school students are arbitrarily divided (even and odd node indexes). A modified contact network is then generated to represent when cohort A is onsite, and one is generated to represent when cohort B is onsite. Each cohort network removes all edges from offsite students, except for their household connections, while maintaining the edges of onsite students. These networks are alternated according to the given cohorting schedule, as applicable.

The degree-based pairwise transmissibility factors $\delta^{(ji)}$ (See Appendix A.1.4.2 Local transmission for details) are calculated according to the connectivities of individuals in the baseline, "everyone onsite" network. The same set of factors derived from this baseline are used to calculate the propensities of local transmission at all times (i.e., for all school days, weekend days, and cohorting days), regardless of which cohort or weekend network is being used to define the structure of close contacts. This reflects an assumption that, for example, the interactions between individuals who are on campus don't become more intense under cohorting just because fewer students are on campus.

**Table A.2.4a** Parameters for the generation of primary school contact networks.

| **Parameter** | **Value** | **Additional description** |
|---|---|---|
| Number of grades | 6 (K-5) | |
| Number of classes per grade | 4 | |
| Number of students per class | 20 | |
| Number of teachergroups | 1 | FARZ parameter for teacher/staff layers: Number of network clusters in teacher/staff layer |
| Teacher/staff mean degree | 5 | Average number of connections each teacher/staff makes with other teachers/staff |
| `alpha` | 5 | FARZ parameter for teacher/staff layers: Strength of common neighbor's effect on edge formation (tunes transitivity, clustering) |
| `gamma` | 5 | FARZ parameter for teacher/staff layers: Strength of degree similarity effect on edge formation (tunes assortativity) |

**Table A.2.4b** Degree distribution plots for a representative primary school network and network property statistics averaged across all primary school contact networks used in our analysis.

| Network | Degree distribution | Network properties |
|---|---|---|
| Overall network | 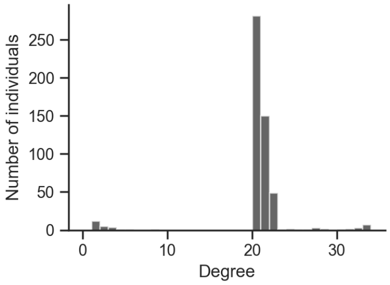 | Degree mean: 20.1<br>Degree std: 4.5<br>Degree $CV^2$: = 0.5<br>Degree assortativity: 0.20<br>Clustering coeff.: 0.91<br>Average path length: 3.6 |
| Student-Student layer | 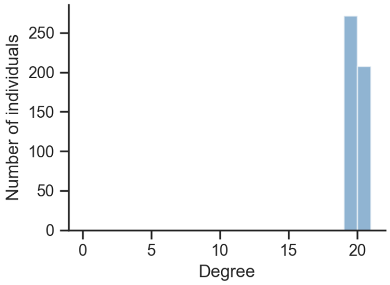 | Degree mean: 19.5<br>Degree std 0.6<br>Degree $CV^2$ = 0.0<br>Degree assortativity: 0.05<br>Clustering coeff.: 0.95<br>Average path length: 3.9 |
| Teacher-Staff layer | 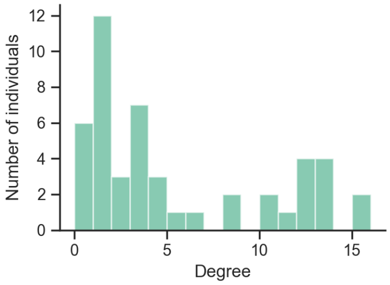 | Degree mean 5.0<br>Degree std 4.8<br>Degree $CV^2$ = 0.8<br>Degree assortativity: 0.46<br>Clustering coeff.: 0.49<br>Average path length: 3.6 |

**Table A.2.4c** Parameters for the generation of secondary school contact networks.

| **Parameter** | **Value** | **Additional description** |
|---|---|---|
| Number of grades | 4 | |
| Number of students per grade | 200 | |
| Number of teachers | 125 | |
| Number of staff | 75 | |
| Number of classes per student | 6 | Number of classes that each student takes each day; i.e., number of teachers with whom each student is connected |
| Percentage of student-teacher grade level matches | 75% | Probability that a student takes a class with, i.e., connects to, a teacher that is associated with their own grade level |
| Number of student social groups per grade | 20 | FARZ parameter for student layers: Number of network clusters in each student layer |
| Student mean intra-grade degree | 16 | Average number of connections each student makes with students in the same grade |
| Student percent inter-grade contacts | 20% | Percent of each student's total student connections that are with students in another grade |
| Number of teacher/staff groups | 10 | FARZ parameter for teacher/staff layers: Number of network clusters |
| Teacher/staff mean degree | 10 | Average number of connections each teacher/staff makes with other teachers/staff |
| `alpha` | 5 | FARZ parameter for all layers: Strength of common neighbor's effect on edge formation (tunes transitivity, clustering) |
| `gamma` | 5 | FARZ parameter for all layers: Strength of degree similarity effect on edge formation (tunes assortativity) |
| `beta` | 0.8 | FARZ parameter for all layers: Probability of edges formation within clusters (strength of cluster structure) |
| `r` | 2 | FARZ parameter for all layers: Maximum number of clusters each node can belong to |
| `q` | 0.5 | FARZ parameter for all layers: Probability of a node belonging to the multiple clusters |

**Table A.2.4d** Degree distribution plots for a representative secondary school network and network property statistics averaged across all primary school contact networks used in our analysis.

| Network | Degree distribution | Network properties |
|---|---|---|
| Overall network | 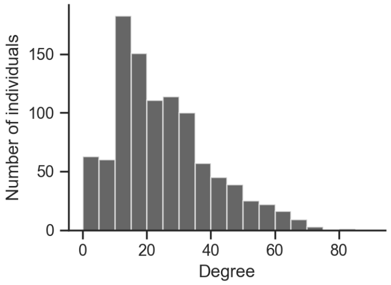 | Degree mean: 24.1<br>Degree std: 15.0<br>Degree $CV^2$: = 0.39<br>Degree assortativity: -0.10<br>Clustering coeff.: 0.16<br>Average path length: 2.6<br><br>References:[38–41] |
| Student-Student layer | 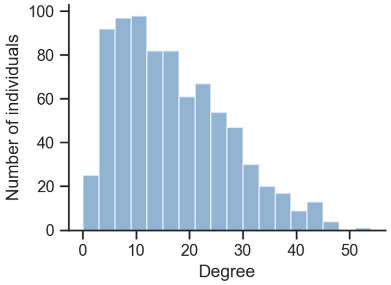 | Degree mean: 16.0<br>Degree std 10.1<br>Degree $CV^2$ = 0.39<br>Degree assortativity: 0.16<br>Clustering coeff.: 0.22<br>Average path length: 2.9<br><br>References:[38–41] |
| Teacher-Staff layer | 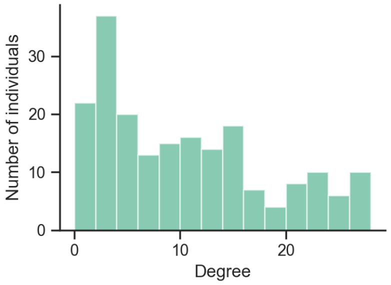 | Degree mean 10.0<br>Degree std 8.2<br>Degree $CV^2$ = 0.64<br>Degree assortativity: 0.39<br>Clustering coeff.: 0.40<br>Average path length: 2.7 |

###### A.2.5 Case Introductions

Exposure to the community is modeled by randomly introducing new cases to the school population according to a Poisson process with an average introduction rate that corresponds to the community prevalence. Each day of the simulation, the number of introductions is drawn from a Poisson distribution using the given introduction rate as the Poisson parameter $\lambda$. Then for each exposure that is to be introduced (if greater than zero), an individual is drawn randomly from the population with selection probabilities proportional to the relative susceptibility of each individual. If the selected individual(s) are susceptible, they become exposed (infected)—otherwise they have been previously infected and their state is left unchanged. This process is handled within the simulation loop adapted from the SEIRS+ Intervention Simulation Loop (Appendix A.2.6.1).

We consider monthly, weekly, and daily introduction rates, as well as single introduction scenarios. These rates roughly correspond to the community prevalences shown in Table A.2.5. These associations between the community prevalence and the rate of introduction to the school population are approximated using the following method. The expected number of new cases to be generated in the overall community is approximated using the equation for the change in the number of infected individuals from the classic SIR model

$$dI_c = \frac{\beta_c S_c I_c}{N_c} = \beta_c S_c \pi_c \,,$$

where $dI_c$ gives the expected number of new infections in the community per day, $N_c$ is the size of the community, $\beta_c$ is the average community transmission rate, $S_c$ is the number of susceptible individuals in the community, and $\pi_c = I_c/N_c$ is the community prevalence (the subscript $c$ denotes a community value). Then the number of these new cases that will land in the school population is assumed to be proportional to the ratio of the size of the school population to the overall community population.

$$\text{expected school introduction rate} = \frac{\beta_c S_c I_c}{N_c}\frac{N}{N_c} \,.$$

When the numbers of current and prior cases in the community ($I_c$ and $R_c$, respectively) are small relative to the size of the community (i.e., $S_c \approx N_c$; this estimation will tend to overestimate the school introduction rate when there is significant susceptible depletion in the community), this can be simplified to a reasonable approximation that does not depend on the size of the overall community

$$\begin{aligned}
\text{expected school introduction rate} &= \frac{\beta_c S_c I_c}{N_c}\frac{N}{N_c} \\
&= \frac{\beta_c (N_c - I_c - R_c) I_c}{N_c}\frac{N}{N_c} \\
&\approx \frac{\beta_c N_c I_c}{N_c}\frac{N}{N_c} \\
&= \frac{\beta_c I_c}{N_c} N \\
&= \beta_c \pi_c N \,.
\end{aligned}$$

Thus the expected rate of introductions to the school population approximately equal to the product of the community transmission rate $\beta_c$, the community prevalence $\pi_c$, and the size of the school population $N$. The community transmission rate is equal to the effective reproduction number $R_{\text{eff}}$ for the community divided by the average infectious period of the disease. Given estimates for these values, the introduction rate can be estimated. This method was used to estimate introduction rates for primary schools ($N$=528) and secondary schools ($N$=1,000) for $R_{\text{eff}}$ in the range (1.0, 2.0), a mean infectious period of 6.2 days, and a range of community prevalence values. The community prevalence ranges for each introduction rate listed in Table A.2.5 are those prevalences for which the expected number of new cases per day in the school population is approximately equal to the listed introduction rate (monthly, weekly, or daily) for some $R_{\text{eff}}$ in (1.0, 2.0) using this method and these parameters.

| | Corresponding Community Prevalence | |
|---|---|---|
| Introduction rate (Poisson $\lambda$) | Primary school (528 individuals) | Secondary school (1000 individuals) |
| Monthly ($\lambda$ = 1/30) | 0.02 - 0.04% | 0.01 - 0.02% |
| Weekly ($\lambda$ = 1/7) | 0.08 - 0.17% | 0.04 - 0.09% |
| Daily ($\lambda$ = 1) | 0.6 - 1.2% | 0.3 - 0.6% |

*For $R_{eff}$ in the 1.0–2.0 range.*

**Table A.2.5 Introduction rates and community prevalences.** Given community transmission in the range of $R_{\text{eff}}$ = 1.0–2.0, this table relates the prevalence of disease in the community to the frequency at which new cases are introduced into a school. Details of how these ranges are estimated are provided in Appendix A.2.5.

##### A.2.6 Interventions

We model several interventions for mitigating the spread of SARS-CoV-2. The SEIRS+ framework provides code for a simulation loop that can implement several interventions, including testing, tracing, and isolation. We make use of a subset of the features in this simulation loop (with minor modification) to implement the mitigation strategies studied in this work.

###### A.2.6.1 Simulation loop

The simulation loop repeatedly calls a function that iterates the Gillespie dynamics of the Extended SEIR Network Model, which determines the next compartment transition (transmission event or disease progression) that will take place, advances the simulation time to the time of that event, and executes the state update. Every time the simulation time crosses an integer value (i.e., a new day is reached), the simulation loop interfaces with the model and its nodes, states, and parameters to implement various intervention procedures. If the Gillespie time to the next event is greater than a day, the simulation advances by a maximum time step that is a fraction of a day to ensure that intervention days are not skipped or irregularly timed. The simulation loop performs the following updates each iteration:

1. Advance the Gillespie compartment transition dynamics
2. If a new day has been reached, execute the following; else Return to (1):
   (a) Update active contact networks and parameters according to the weekend and cohorting schedule when applicable (see Appendix A.2.6.3 Cohorting and Appendix A.2.4.5 Cohort contact networks).

(b) Introduce new community exposure cases (see Appendix A.2.5 Case Introductions).

(c) Isolate symptomatic individuals who are compliant with self-isolation upon symptom onset (see Appendix A.2.6.4 Isolation).

(d) If the current day is part of the testing cadence, test individuals who are eligible for testing when applicable (see Appendix A.2.6.2 Testing); else skip.

(e) Isolate individuals who have received a positive test result (following the test result lag time) and their classmates when applicable (see Appendix A.2.6.4 Isolation)

(f) Return to (1)

More details about these interventions are provided in the following sections.

###### A.2.6.2 Testing

We consider proactive testing that is executed according to one of several testing cadences (including no testing) shown in Table A.2.6. These cadences define which groups of individuals are tested and on which days of the week. On a designated testing day, all individuals who are eligible to be in the testing pool are tested. An individual is considered part of the testing pool when they:

- Are a member of one of the groups designated in the testing cadence
- Are not currently in isolation
- Have not already had a positive test result
- Have not already recovered from the disease
- Have not been vaccinated
- Are compliant with testing

We assume that 100% of teachers and staff are compliant with testing, but 25% of students are non-compliant and thus never get tested. Students are assigned a compliance status randomly according to this probability when the model is initialized.

We model realistic temporal test sensitivities consistent with RNA-based tests. We assume 0% sensitivity for individuals in the exposed (latent) state. We assume 75% sensitivity for individuals in the first 2 days of their pre-symptomatic period and 80% sensitivity for any pre-symptomatic days beyond that. Sensitivities for symptomatic

| **Proactive Testing Cadence** | Mon | Tue | Wed | Thu | Fri | Sat | Sun |
|---|---|---|---|---|---|---|---|
| No Testing | | | | | | | |
| Teachers Weekly | 🟢🟡 | | | | | | |
| Teachers Semiweekly | 🟢🟡 | | | 🟢🟡 | | | |
| Everyone Weekly | 🟢🟡🔵 | | | | | | |
| Everyone Semiweekly | 🟢🟡🔵 | | | 🟢🟡🔵 | | | |

🟢 Teachers 🟡 Staff 🔵 Students

**Table A.2.6 School testing cadences.** We explore the consequences of five testing cadences, from no testing to testing all members of the school community twice a week. Once-weekly testing takes place every Monday, and Semiweekly testing takes place on Mondays and Thursdays.

and asymptomatic individuals alike follow the time course shown in Figure A2, which follows data for RNA-based tests from Levine-Tiefenbrun et al.[82] We assume there are no false positives. We evaluate the sensitivity of model outcomes to the testing method in Appendix B.6, where we consider test sensitivities in line with data for antigen-based tests.

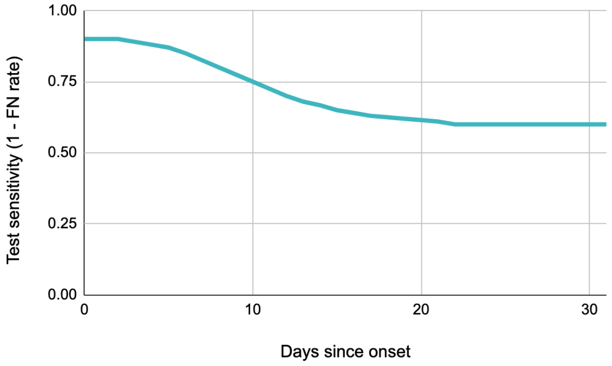

**Figure A2: Test sensitivities.** The probability of returning a positive test results when testing an individual in a symptomatic ($I_{\text{sym}}$ or asymptomatic $I_{\text{asym}}$ infectious state as a function of the number of days since entering that state (i.e., days since onset of symptoms). The test sensitivity is equivalent to 1 minus the false negative rate.

There is a 1 day lag (exactly 24 hours) between administering a test and receiving the result. Individuals that receive a positive result enter isolation (i.e., move into a quarantine compartment) immediately upon receiving the positive result, thus 1 day after being tested. We assume that all individuals in the school population are compliant with entering isolation upon a positive test result.

###### A.2.6.3 Cohorting

Cohorting consists of dividing students into two groups and following an alternating schedule in which only one group is on campus at a time. We consider two cohorting schedules, one where the group of students that is onsite alternates daily, and one where the onsite group alternates weekly (in addition to no cohorting). The cohorting schedules are summarized in Figure A3.

| **Cohort Schedule** | | Mon | Tue | Wed | Thu | Fri | Sat | Sun |
|---|---|---|---|---|---|---|---|---|
| All Students Onsite | Week 1 | AB | AB | AB | AB | AB | | |
| | Week 2 | AB | AB | AB | AB | AB | | |
| Alternate A/B Cohorts Daily | Week 1 | A | B | A | B | A | | |
| | Week 2 | B | A | B | A | B | | |
| Alternate A/B Cohorts Weekly | Week 1 | A | A | A | A | A | | |
| | Week 2 | B | B | B | B | B | | |

A Cohort A on campus   B Cohort B on campus

**Figure A3**

Cohorting is implemented by alternating between different versions of the contact network in which one group of students or the other has their connections with the school population removed, except for any connections to individuals in their own household. Global transmission remains active for students who are offsite due to the cohort schedule, which can be thought of as students having some propensity to interact with other members of the school population outside of school. We assume that all students comply with the cohorting schedule. Students who are offsite due to cohorting are not considered to be in isolation, and these students are still part of the testing pool when otherwise applicable.

###### A.2.6.4 Isolation

When an individual enters isolation, they are moved into the quarantine compartment that corresponds to their disease state at the time of isolation. This compartment transition is executed "manually" by the simulation loop, separate from the Gillespie or residence time-based transition dynamics (Appendix A.1.3 Dynamics). While in isolation, individuals transition between quarantine compartments according to the same disease state residence times that are used when not in isolation. The set of close contacts for isolated individuals is given by a distinct quarantine contact network, which includes connections to members of the quarantined individual's household but no other members of the school population. In addition, quarantined individuals make no casual contacts (i.e., no global transmission). Isolated individuals remain in the quarantine sequence of compartments until their total isolation period has elapsed, at which time they are moved into the non-quarantine compartment that corresponds to their current disease state. We use a 10 day isolation period for all individuals, following the current CDC recommendation.[93]

In this model, individuals may enter isolation upon the onset of symptoms (if compliant), after receiving a positive test result, or when another member of their classroom has tested positive (for primary schools, when applicable).

We assume that 20% of all individuals elect to self-isolate upon the onset of symptoms. The symptomatic isolation compliance status of individuals is assigned randomly according to this probability when the model is

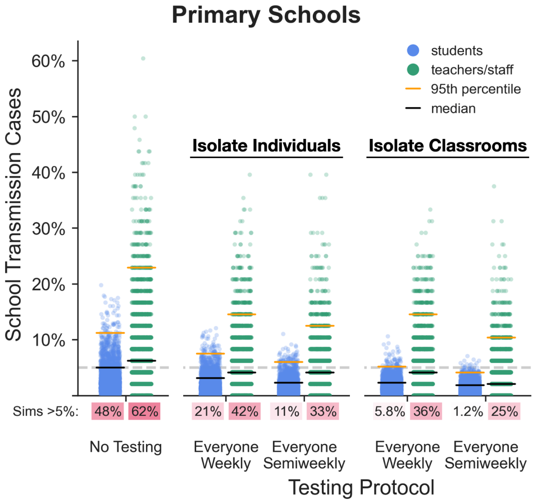

**Figure A4: Effect of isolating classrooms.** We consider two quarantine strategies for primary schools: (1) isolate single individuals who receive a positive test result, and (2) isolate the entire classroom (students and teacher) associated with an individual who receives a positive test. The distributions of school transmission cases as a percentage of the school population are shown for 1,000 simulations of each testing and isolation strategy with $R_0$=1.5 and approximately weekly new case introductions. Under each jitter distribution we list the percentage of simulations that result in outbreaks affecting more than 5% of the population. Black and orange lines represent median and 95th percentile outcomes respectively.

initialized. For compliant individuals, there is a 1 day lag between transitioning into the symptomatic compartment and entering isolation. Individuals who are asymptomatic and thus enter the asymptomatic compartment rather than the symptomatic compartment never self-isolate. Symptomatic self-isolation does not trigger isolation of the symptomatic individual's contacts. Note that the rate of asymptomatic disease is assumed to be different in primary school-aged children (40%) and adults/adolescents (30%) while the rate of symptomatic isolation compliance is constant, so the effective rate of symptomatic self-isolation is lower in young children.

In primary schools, where classroom groupings are stable, we also consider scenarios where entire classrooms (i.e., all students and the teacher) isolate when any one member of the classroom tests positive. In such a case, the entire group enters isolation at the same time immediately after the index case receives their test result (i.e., 1 day after being tested).

###### A.2.6.5 Vaccination

We also evaluate the effectiveness of vaccinating teachers and staff on mitigating transmission in schools. We use the following working definitions with regard to vaccination in this model:

- Uptake: The percentage of individuals who receive a vaccine
- Effectiveness: The percentage of individuals receiving the vaccine that have an efficacious response. An efficacious response is characterized by an immune response that protects the vaccinated individual from falling ill and prevents that individual from transmitting disease.
- Reduction in transmissibility: The factor by which individual transmissibility is reduced for individuals with an efficacious response to vaccination

We model the scenario where 100% of teachers and staff are vaccinated with a vaccine that has 90% effectiveness (and no students are vaccinated). As noted above, an efficacious response also blocks 100% of transmission (i.e., the vaccinated individual's transmissibility is reduced to 0). The individuals that are to have an effective response are chosen randomly according to the probability of effectiveness when the model is initialized. Teachers are vaccinated and individuals with effective responses have their transmissibilities reduced before the simulation time begins. Individuals with effective immune responses are not included in case counts due to their immunity to the disease. Individuals with ineffective responses have no change in transmissibility or other parameters, can still contract and transmit the virus, and are included in case counts.

### Appendix B Sensitivity of Outcomes to Model Parameters

School administrators and public health officials need ways of evaluating and mitigating the risk of disease transmission in school populations. Their challenge is exacerbated by ongoing uncertainty about disease characteristics, changing transmission and behavioral conditions, and variability from one population to the next. Even within a relatively circumscribed population such as a school, many variables influence the likelihood and severity of outbreaks. One benefit of a mechanistic modeling study is that the relative influence of parameters can be explored systematically.

In the main text we focus on two specific models intended to capture the dynamics of disease transmission within primary and secondary school populations, respectively. These models are characterized by the disease course, transmission, and contact network parameterizations described in detail in Appendix A.2. Our choices of parameters in those models are data driven, where quality data are available, and are intended to capture the general features that are of epidemiological relevance for primary and secondary school populations; see Appendix A.2 for more information. In the real world, no two schools are alike, and few if any will exactly match our choice of parameters. In this section, we explore the effects of changes in parameter values on epidemic outcomes. This analysis provides information about the sensitivity of the model to various parameters, and offers insights about which parameters may merit special attention from pandemic response planners.

In each of the following sections, we present a sensitivity analysis across one or two referenced parameters at a time, while all other parameters are held at their baseline values as outlined in Table A.2.1.

**Readers can also explore the impacts of mitigation strategies and the effects of various parameters on model outcomes using an interactive web application at https://www.color.com/return-to-school-model.**

#### B.1 Transmissibility

##### B.1.1 Basic reproduction number $R_0$

The basic reproduction number $R_0$ is a critical parameter of disease dynamics. In our model, each individual $i$ is assigned an *individual reproduction number* $R_0^{(i)}$, which is the expected number of secondary cases that the individual generates when infectious in a fully susceptible population (Appendix A.1.4). The *basic reproduction number* $R_0$ is the average number of secondary cases to be expected from an infectious individual in a wholly susceptible population. Any case has the potential to initiate a chain of transmission. There is stochasticity and individual variation associated with transmission events, but the greater the basic reproduction number the greater the number of secondary cases per infected individual on average—and the more likely it is that ongoing transmission occurs. In addition, the higher the value of $R_0$, the more likely that any single introduction seeds an outbreak, and the higher is the herd immunity threshold.

Figure B1 and Figure B2 show the average total number of school transmission cases and the mean cluster size

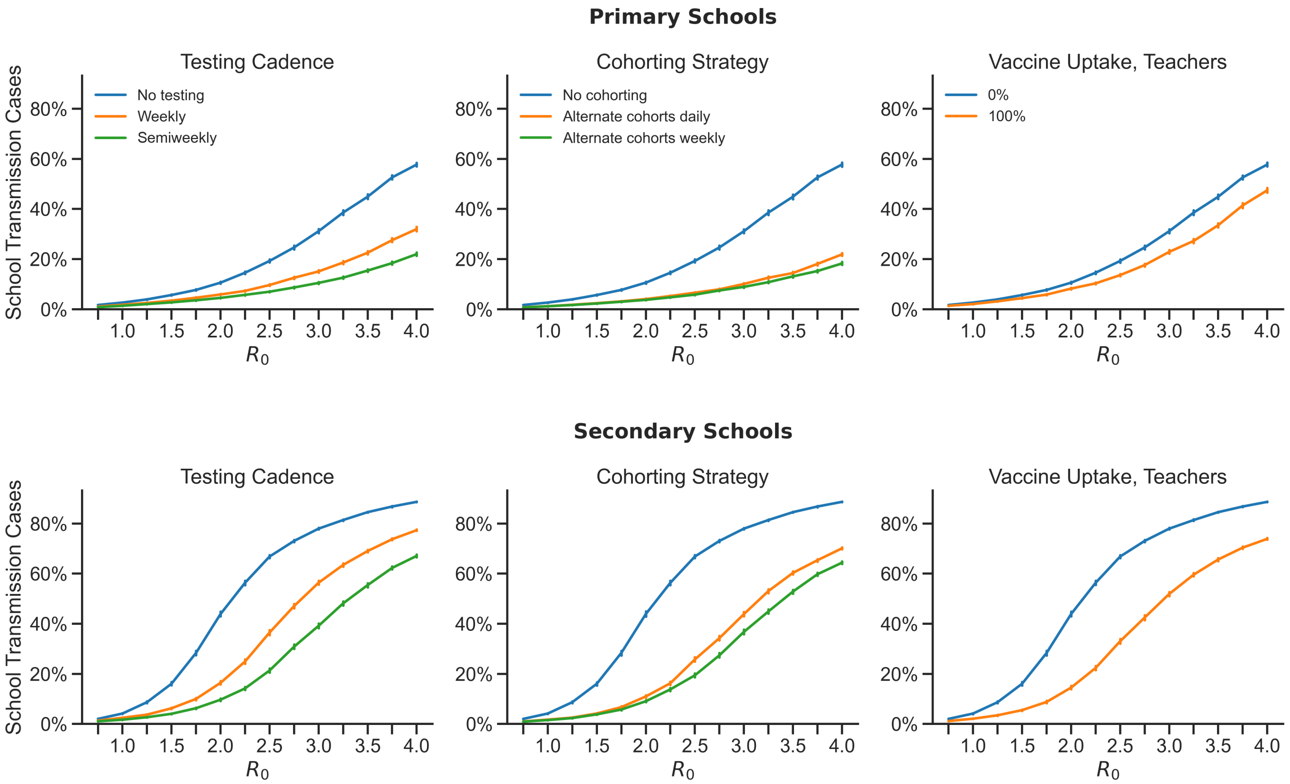

**Figure B1: Cases increase dramatically with increasing basic transmission number $\mathbf{R_0}$.** The average number of school transmission cases are shown as a percentage of the total school population for $R_0$ values from 0.75 to 4.0. In this figure and in those that follow throughout this appendix, cases are introduced into the school once a week on average, and baseline values are used for all other parameters as outlined in Table A.2.1. Error bars give the 95% confidence interval around the mean for 1000 replicates. Outcomes are shown for primary schools (top) and secondary schools (bottom). As a baseline we consider scenarios with no mitigation (blue lines), and to this we compare weekly or semiweekly testing of students and teachers (left column), alternating student cohorts on a weekly or daily schedule (center column), and vaccination of teachers (right column).

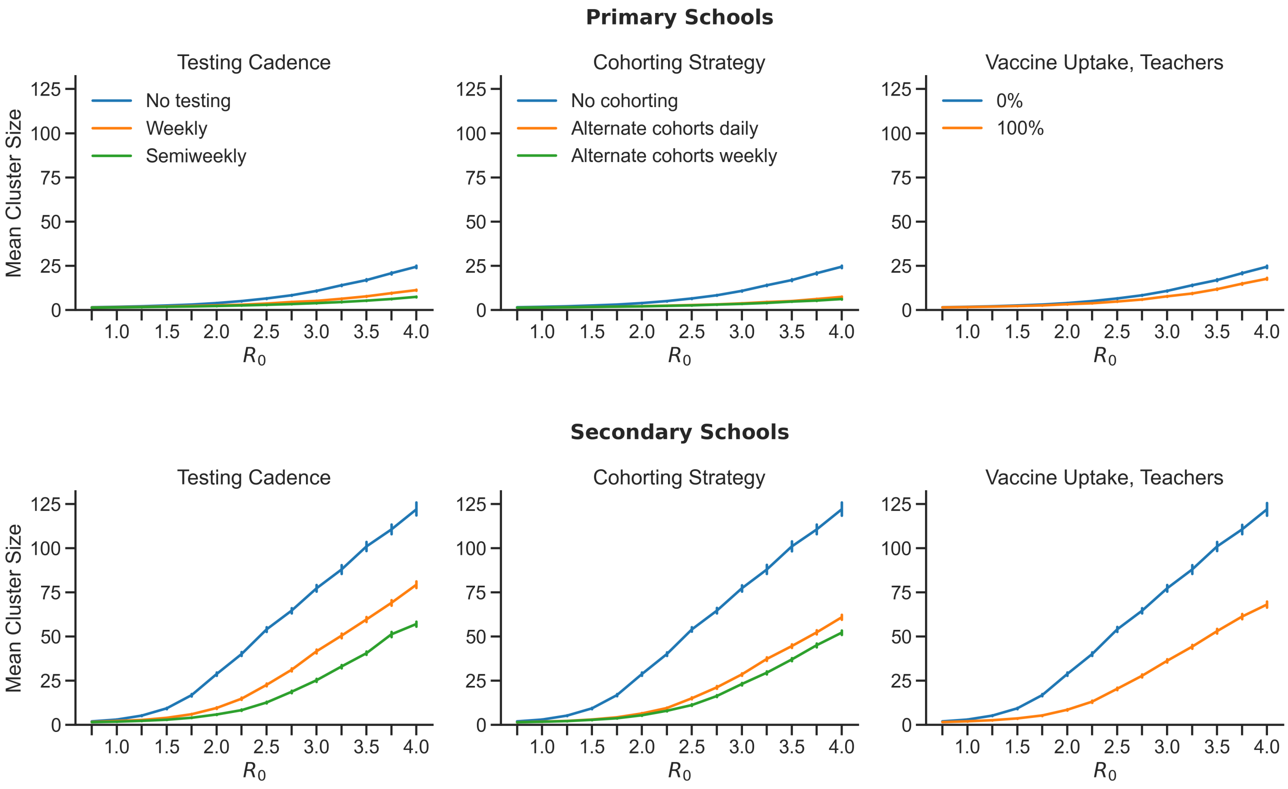

**Figure B2: Average cluster size increases with increasing basic reproductive number** $R_0$. The average cluster sizes per introduction in primary schools (top) and secondary schools (bottom) are shown for a range of $R_0$ values.

per introduction, respectively, for primary and secondary schools across a range of $R_0$ values. As $R_0$ increases, the mean cluster size increases and the average number of school transmission cases increases as well. For a given $R_0$, primary schools have smaller clusters and fewer total school transmission cases on average, which is due in large part to the decreased susceptibility of primary school-aged children (Appendix B.3.1). The relative benefits of mitigations are seen in the differences between curves in the same plot. All mitigations are beneficial even at low values of $R_0$, and the absolute benefits of all mitigations increase along with $R_0$. While the marginal benefits of testing semiweekly rather than weekly testing and of alternating cohorts on a daily rather than weekly basis are relatively small when $R_0$ is low, these advantages become sizeable at higher levels of $R_0$.

The value of $R_0$ in a given population depends on many factors that influence the likelihood of transmission, including but not limited to individual behavior, interaction patterns, inherent transmissibilities of strains in the population, and features of the school environment. Of the parameters considered in our model, changes in $R_0$ have the greatest marginal effects on outcomes. This can make quantitative prediction difficult, because $R_0$ is seldom known precisely and is subject to fluctuations over time. But the qualitative trends are consistent: everything gets worse with higher $R_0$ values, and planners should make every effort to keep $R_0$ to a minimum using control measures such as physical distancing, mask wearing, and proper ventilation. Proactive mitigation strategies such as testing, cohorting, and vaccinating teachers can reduce the effective reproduction number further, but these strategies cannot sufficiently mitigate risk if the baseline level of transmission is too high.

##### B.1.2 Overdispersion of individual transmissibility

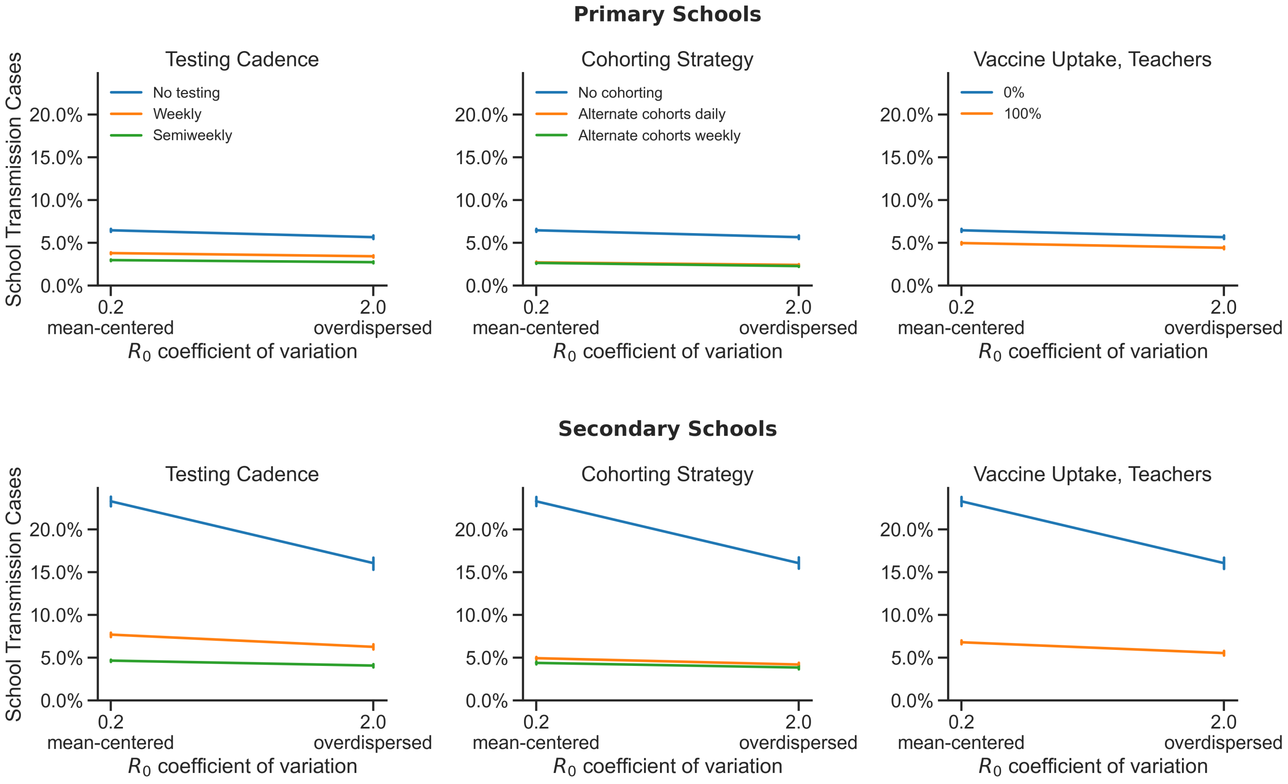

**Figure B3: Variation in individual transmissibility has at most a modest effect on school transmission.** The average number of school transmission cases are shown as a percentage of the total school population for mean-centered (gamma coeff. of variation = 0.2) and overdispersed (gamma coeff. of variation = 2.0) distributions of individual reproduction numbers. In primary schools under all conditions and in secondary schools with any testing, cohorting, or vaccination in place, variation in individual transmissibility has no effect on trasmission in schools. In secondary schools with no mitigations in place, overdispersed transmissibilities reduce the mean transmission somewhat.

In our model, individuals vary in their propensity to transmit disease. Each individual is assigned an individual reproduction number $R_0^{(i)}$ (i.e., the expected number of secondary cases that the $i$th individual would generate when infectious in a fully susceptible population). Individual reproduction numbers are drawn from a gamma distribution with a mean equal to the baseline $R_0$ and a shape parameterized by a coefficient of variation. In main body of this paper we assume that individual transmissibilities follow an overdispersed (long-tailed) distribution, with approximately 20% of individuals contributing 80% of the total expected number of secondary cases (the "80/20 rule").[27–29] Under such a distribution, the majority of individuals are expected to generate fewer than one secondary case while a minority of individuals are expected to contribute many cases. Evidence from contact tracing studies suggest that transmission of SARS-CoV-2 follows this 80/20 rule,[30–34] but we might also consider cases with reduced variation in individual transmissibility, such that most individuals are expected to generate a number of cases close to the value of $R_0$. A gamma distribution with a coefficient of variation of $2.0$ gives an overdispersed distribution where 80% of individuals fall in the upper 20th percentile of the distribution. A gamma distribution with a coefficient of variation of $0.2$ gives a bell-shaped, mean-centered distribution with less variation in transmissibility.

Figure B3 shows the effect of the amount of variation in individual transmissibility on the number of transmissions in school populations. Populations with overdispersed distributions of individual transmissibilities exhibit fewer school transmission cases in principle,[29,94,95] as is especially apparent in secondary schools without mitigation (blue lines in bottom row). In overdispersed populations, a small number of individuals may generate a large number of cases should they become infected, but a majority of the population have individual reproduction numbers $R_0^{(i)} < 1$ and are expected to contribute less than a single secondary case on average. Some outbreaks that go through highly-transmissible individuals are large, but the average transmission chain is short lived. On the other hand, most individuals in a population with a mean-centered distribution of individual transmissibility are expected to generate at least one secondary case if the mean reproduction number $R_0$ is greater than 1. Super spreading is less common, but the average number of transmissions is higher. We also see in Figure B3 that once mitigations are introduced, observed outcomes are insensitive to the amount of variation in individual transmissibility. This is presumably because when transmission chains are broken early, individual variation in transmissibility does not come much into play.

##### B.1.3 Pre-symptomatic and asymptomatic infectiousness

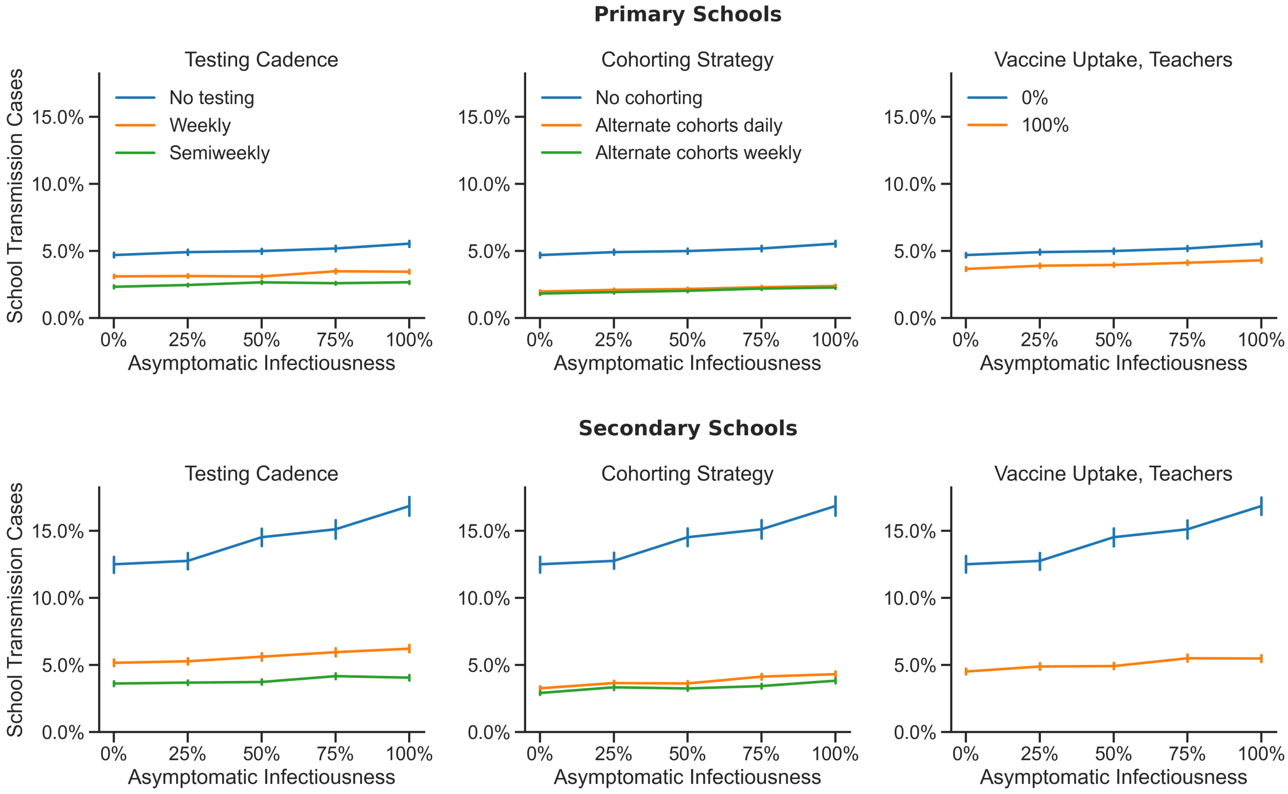

**Figure B4: Increasing asymptomatic transmissibility drives increased transmission in secondary schools when interventions are not used, but has minimal effects under other other conditions** The average number of school transmission cases are shown as a percentage of the total school population for a range of asymptomatic transmissibily values, relative to the transmissibility of symptomatic individuals.

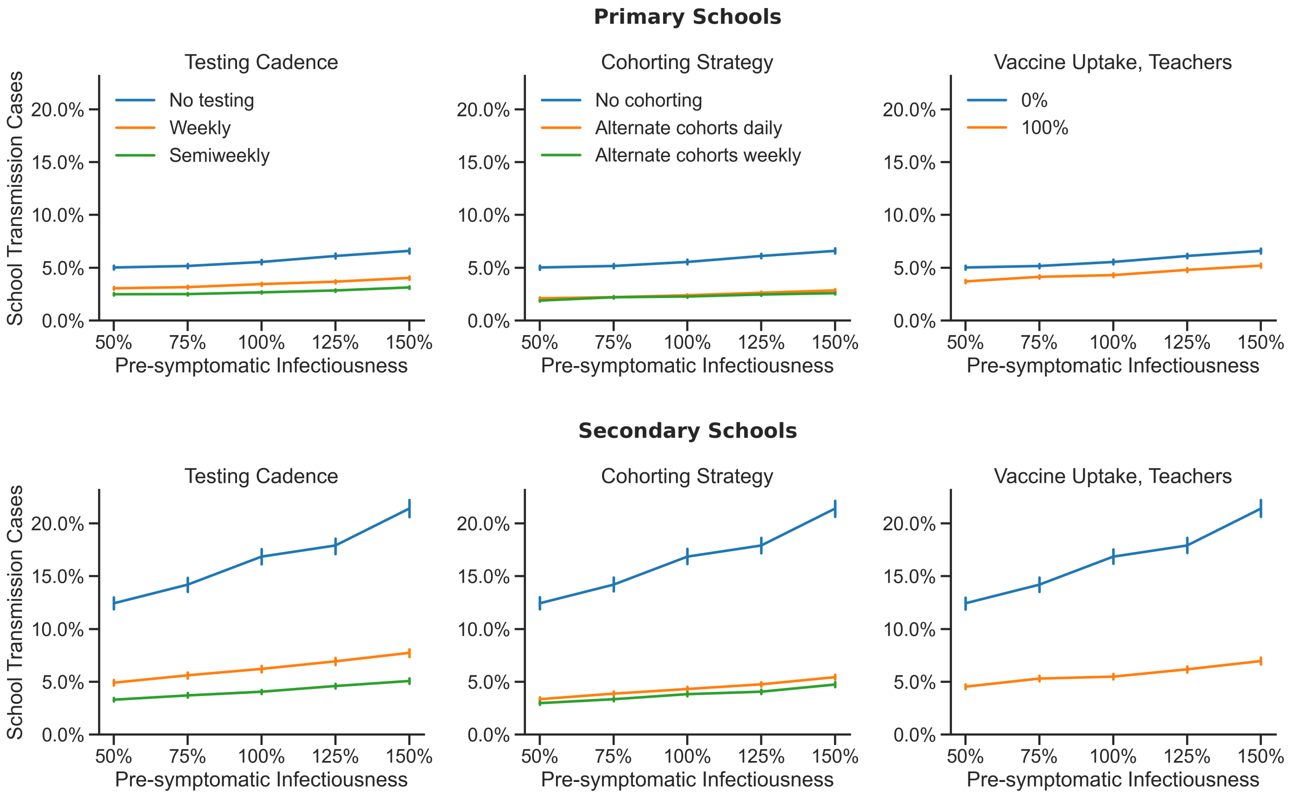

**Figure B5: Increasing presymptomatic transmissibility drives increased transmission in secondary schools when interventions are not used, but has modest effects under other other conditions.** The average number of school transmission cases are shown as a percentage of the total school population for a range of pre-symptomatic transmissibility values relative to the transmissibility of symptomatic individuals.

Some studies suggest that pre-symptomatic individuals may contribute a disproportionately high number of cases relative to the duration of this disease state, and asymptomatic individuals may contribute disproportionately few cases relative to symptomatic individuals[73–75]. However, these studies draw on contact tracing data (e.g., incidence rates and attack rates associated with individuals in different disease categories), and it is unclear how much of the differences in transmission between pre-symptomatic, symptomatic, and asymptomatic individuals are attributable to differences in biological transmissibility (e.g., viral load and shedding) as opposed to differences in behavior or other factors. Without more data regarding the relative viral loads and viral shedding of pre-symptomatic and asymptomatic individuals, we opt to assume that there is no difference in biological transmissibility between the pre-symptomatic, symptomatic, and asymptomatic states. This assumption is conservative with respect to asymptomatic transmission in the sense that our results will err on the side of overestimating the number of school transmission events if asymptomatic or pre-symptomatic individuals are indeed less infectious than symptomatic individuals. However, our results may overstate the relative benefit of mitigations in such a case. If pre-symptomatic individuals are more infectious than symptomatic individuals, our results may underestimate the amount of transmission that is likely to occur in schools, but the importance of proactive mitigations will be even greater.

Figure B4 shows the average number of school transmission cases for primary and secondary schools across a

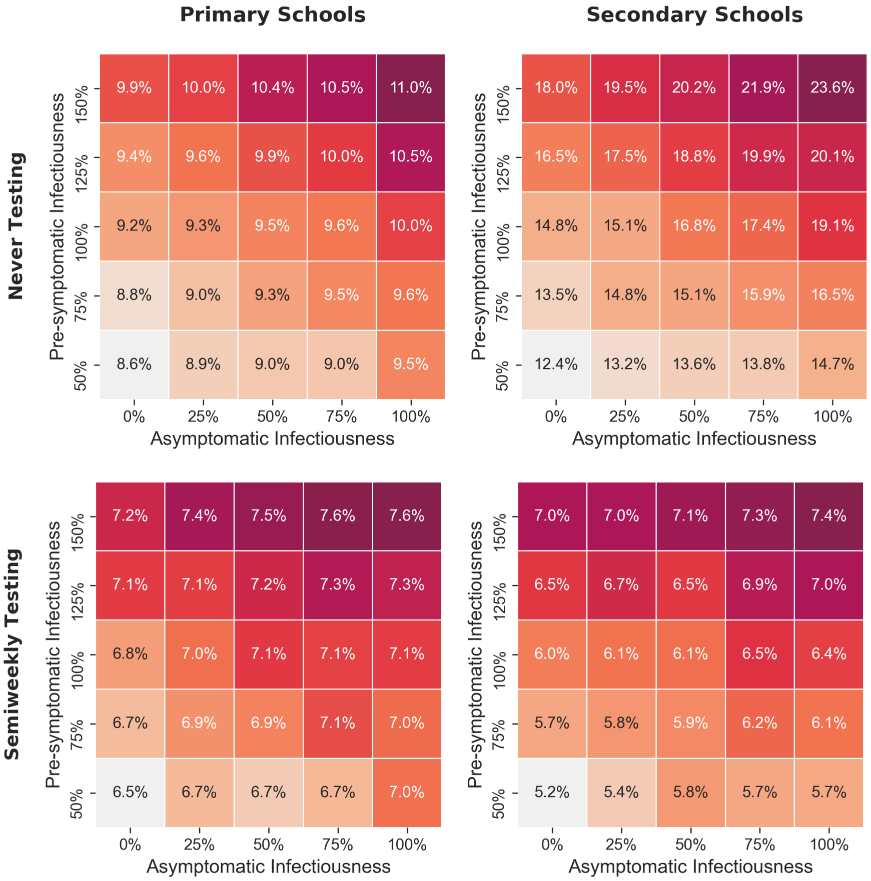

**Figure B6: Increasing asymptomatic and presymptomatic trasnsmissibilities compound to increase overall transmission.** The heatmap cells give the average number of school transmission cases shown as a percentage of the total school population for combinations of pre-symptomatic and asymptomatic infectiousness relative to symptomatic infectiousness. Outcomes are shown for primary schools (left column) and secondary schools (right column) in scenarios with no testing (top row) and semiweekly testing (bottom row). Cases are introduced into the school once a week on average, no other mitigations are in place, and baseline values are used for all other parameters as outlined in Table A.2.1. The color maps of each heatmap have been independently scaled to highlight within-treatment sensitivity to pre-symptomatic and asymptomatic infectiousness.

range of asymptomatic transmissibilities relative to symptomatic cases. Particularly in the absence of interventions, the average outbreak size is reduced somewhat when asymptomatic cases have reduced transmissibility. This effect is more pronounced in secondary schools than in primary schools. The marginal benefits of mitigations are greater at higher levels of asymptomatic infectiousness, but there is still significant benefit to all interventions in both settings even when asymptomatic individuals are not infectious at all.

Figure B5 shows the effect of varying pre-symptomatic infectiousness. As with asymptomatic infectiousness, the average number of school transmission cases and the benefits of mitigations are greater at higher levels of pre-symptomatic transmissibility. If pre-symptomatic individuals are indeed more infectious than symptomatic individuals (independent of changes in behavior, etc.), then proactive interventions such as testing, cohorting, and vaccination have even greater value.

Figure B6 illustrates the effect of varying pre-symptomatic and asymptomatic infectiousness together. In the absence of testing or other mitigations (top), model outcomes are approximately equally sensitive to changes in either parameter, although changes in asymptomatic infectiousness have a slightly greater impact. The effect of increasing pre-symptomatic infectiousness is approximately offset by a commensurate decrease in asymptomatic infectiousness. When semiweekly testing is implemented, many asymptomatic cases are detected and isolated, and outcomes become insensitive to the level of asymptomatic infectiousness. However, outbreak size is still sensitive to pre-symptomatic infectiousness even with frequent testing, because some individuals will not be tested during their presymptomatic period and because testing will fail to detect some cases within the pre-symptomatic period due to the false negative rate of the test being higher during this period.

#### B.2 Population mixing (global transmission)

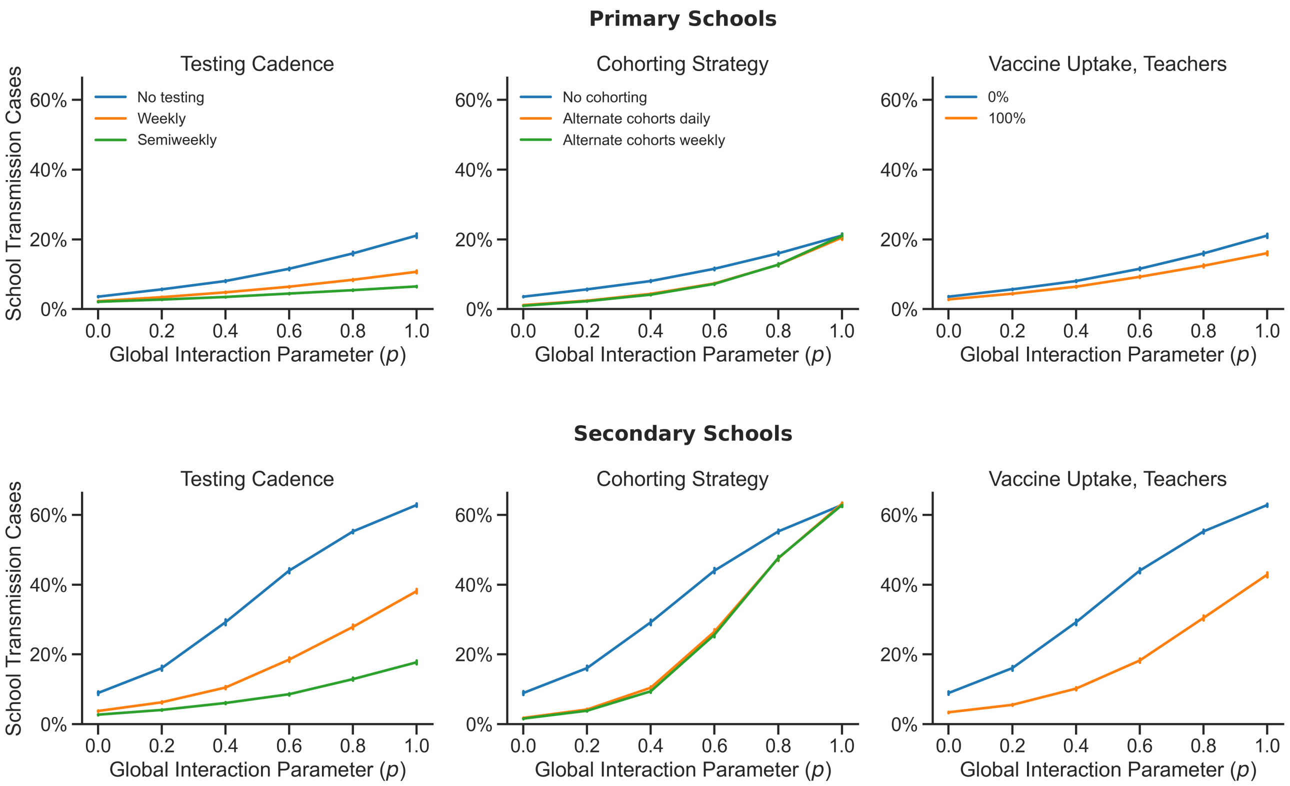

**Figure B7: Case increase as transmission shifts from primarily along the contact network to more globally among weak contacts.** The average number of school transmission cases are shown as a percentage of the total school population for a range of global transmission values.

Disease transmission may occur either from close contacts (local transmission) defined by the contact network structure or from casual contacts (global transmission). Close contacts are individuals with whom one has repeated, sustained, or close proximity interactions on a regular basis, while casual contacts are individuals with whom one has incidental, brief, or superficial contact. Casual contacts are assumed to be randomly sampled from the population as in a well-mixed population. In the context of a school population, a student's close contacts might include their teacher(s), other students in their class(es) or activities, their friends, and their siblings. Global transmission may represent transmission among students, teachers, or staff that interact incidentally in class, in the cafeteria, in the hall, on the bus, etc. A network locality parameter $p$ sets the relative weight of interaction among casual (global) contacts in the population. This parameter can be seen as setting the amount of mixing in the population, where higher values of $p$—more global interaction—correspond to more random encounters among individuals throughout the population. In the paper we assume that 80% of transmission occurs between close contacts on the contact network and 20% of transmission occurs globally (i.e., $p = 0.2$).

Figure B7 shows the average number of school transmission cases for primary and secondary schools across a range of global interaction weights, $p$. When $p = 0$ there is no global interaction, and all transmission occurs between close contacts on the contact network. As $p$ increases, the population becomes increasingly mixed as more and more transmission occurs globally rather than on the network. When $p = 1$ the population is well-mixed, and the dynamics of transmission reduce to a stochastic implementation of a standard mass action compartment model. As the population becomes more mixed, the effect of stochasticity on the fates of transmission chains decreases, and the probability increases that a single case introduction will seed an outbreak that proceeds all the way to the herd immunity threshold. Since the influence of the contact network is diminished in a relatively mixed population, the efficacy of cohorting, which is defined as a partitioning of close contacts among students, decreases as the extent of global interaction increases.

These results suggest that schools should make efforts to limit the amount of mixing and opportunities for global transmission in their populations. This might include restructuring lunch periods, passing periods, transportation logistics, etc. in order to limit congregation and interaction among groups of otherwise "unconnected" individuals at school.

#### B.3 Differences between primary and secondary schools

Across parameter ranges and interventions, we find that compared to secondary schools, primary schools consistently have fewer school transmission cases and lower probabilities of sizable outbreaks. In our model, the key characteristics that differentiate primary and secondary schools are (1) the susceptibility of students and the (2) the sturcture of contact networks. In this section we investigate the relative contribution of these factors to the differences in outcomes across these settings.

##### B.3.1 Susceptibility of students

A fundamental difference between primary and secondary school populations is the age of the students. Evidence suggests that primary school-aged children are less susceptible to COVID-19 than adults, while secondary school-aged adolescents are approximately equally susceptible to adults.[16,20] In the main text, we have assumed that primary school students are 60% as susceptible as adults, and secondary school students are 100% as susceptible. Given the lower susceptibility of the majority of individuals in primary school populations, we would expect primary schools to have fewer and smaller outbreaks. This is borne out in our model as well as in empirical data.[16–19]

Figure B8 shows the effect of varying student susceptibility from 60% to 100% that of adults in both primary and secondary schools. We see that given that same level of student susceptibility, primary and secondary schools have similar outcomes and show similar relative benefits to mitigation measures. When primary school students are 100% as susceptible as adults, outcomes in primary schools closely resemble those of secondary schools. When the susceptibility of secondary school students is reduced to 60% that of adults, the outcomes in secondary schools resemble those of primary schools. These patterns suggest that student susceptibility is a key determinant of the

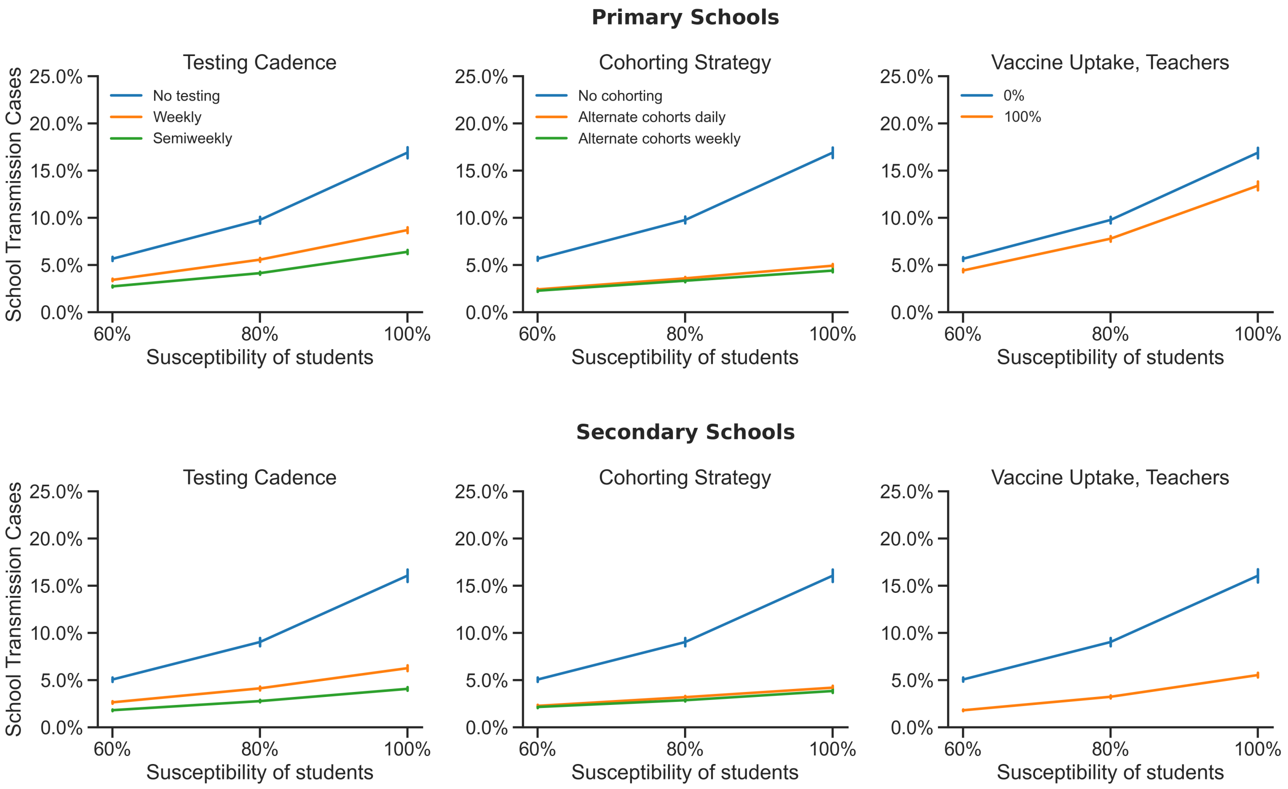

**Figure B8: When primary and secondary school students are equally susceptible to infection, primary schools and secondary schools exhibit similar case rates.** The average number of school transmission cases are shown as a percentage of the total school population, for a range of student susceptibilities relative to adults, using primary school (upper row) and secondary school (lower row) network structures. Baseline values are used for all other parameters as outlined in Table A.2.1. The strong similarities between the primary and secondary school plots for any given level of susceptibility indicate that the differences in network structure have minimal effects.

epidemiological outcomes in schools. In the next section we will see that outcomes are relatively insensitive to the differences in contact network structure between these two school settings, which supports the conclusion that student susceptibility is the an important driver of epidemiological differences between primary and secondary schools.

##### B.3.2 Modularity of contact networks

Another major difference between primary and secondary schools is the structure of the contact network that represents the patterns of interaction in these populations. Perhaps the most significant difference between these two characteristic contact networks is their modularity. Primary schools are organized into classroom groups that interact closely throughout the day, which represents a highly modular contact structure. In secondary schools, students attend multiple classes each day, teachers interact with multiple groups of students, and students participate in activities outside of class—all of which contribute to a less modular interaction network. Indeed, the contact network used in our primary school model is significantly more modular than the secondary school network (mean clustering coefficients of 0.91 and 0.16, respectively). In general, population modularity tends to dampen the severity of epidemics.[24,25,41] When individuals belong to more insular groups, they have fewer sources of possible exposure

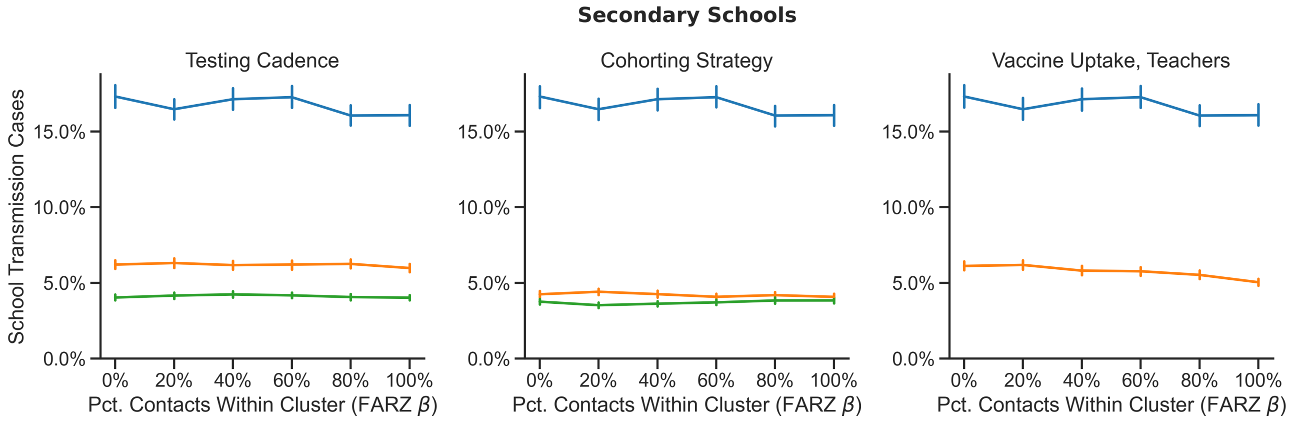

**Figure B9: In secondary schools, transmission is not sensitive to the network structure of within-grade interactions.** In the secondary school model, the FARZ network generator is used to generate a student contact network layer for each grade level. Students are assigned to network clusters, and the FARZ $\beta$ parameter sets the probability of within-cluster edge formation. The average number of school transmission cases are shown as a percentage of the total school population for a range of $\beta$ values, with higher values representing more modular networks.

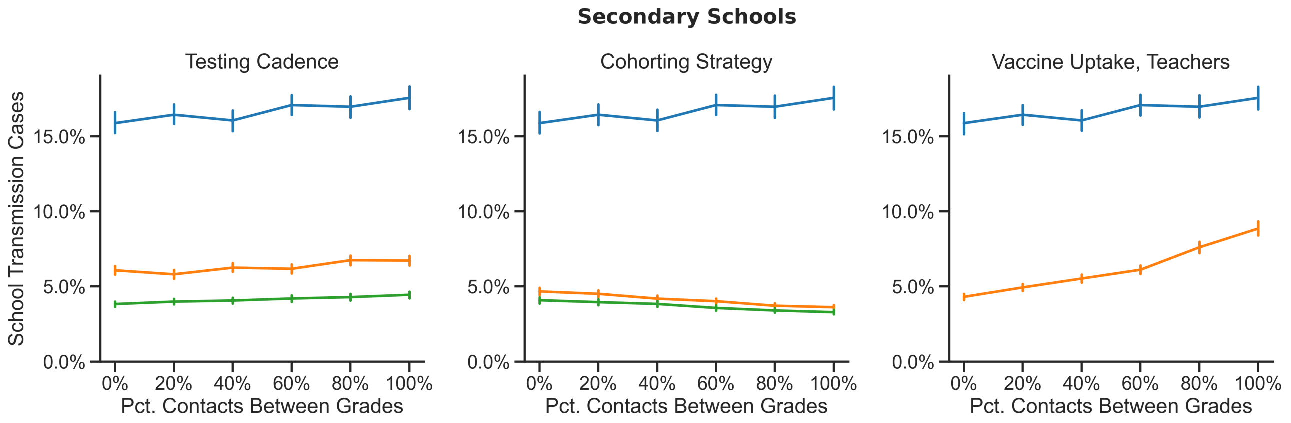

**Figure B10: In secondary schools, transmission is only slightly sensitive to the extent of between-grade interactions.** In the secondary school model, a student contact network layer is generated for each grade level. A parameterized fraction of each student's total number of edges are designated for between-grade interactions. The average number of school transmission cases are shown as a percentage of the total school population for a range of between-grade connectedness values, with lower values representing more modular student contact networks.

when susceptible and a more restricted and more correlated pool of potential infectees when contagious. This expectation is consistent with our findings that primary schools tend to have fewer and less severe outbreaks than secondary schools, but how much of the difference that we observed is attributable to network structure rather than other factors?

To answer this question we probe changes to the secondary school network that make it more modular and thus more similar to the primary school network. The secondary school contact network consists of five interconnected FARZ network layers: a student layer for each grade level and a teachers-and-staff layer (Appendix A.2.4). The FARZ network generator assigns each student to 1-2 network clusters, which may be thought of as groups of students that take the same classes, participate in the same activities, belong to the same social circle, or are otherwise more likely to interact and share contacts with each other than with other students. The FARZ $\beta$ parameter sets the expected fraction of each student's close contacts that are within their own network cluster. Therefore this parameter modulates the modularity of the student layer for each grade level. When $\beta = 1$ students belong to highly modular groups that interact amongst themselves, and when $\beta = 0$ these groups are degenerate as students interact exclusively with students from other groups. The secondary school contact network is also parameterized by the fraction of each student's contacts that are between grade levels. Decreasing the amount of inter-grade interaction makes the overall network more modular, and vice versa. Our baseline secondary school model features a contact network where 80% of each student's contacts are with students in the same grade, and 80% of those within-grade contacts are with students within their own network cluster.

Figure B9 and Figure B10 show the effect of varying these modularity parameters on the average number of school transmission cases in secondary schools. Network modularity is increased by increasing the FARZ $\beta$ parameter or by decreasing the fraction of inter-grade contacts. We find that the model outcomes are largely insensitive to changes in these parameters. This suggests that differences in the modular structure of students' interaction networks are not a substantive driver of the differences in outcomes in primary and secondary schools. Even when the student-student contact network is highly modular, other forms of interaction forge transmission chains across groups, including teacher-student interactions, household transmission, and global transmission due to incidental interactions. We have previously seen that model outcomes are highly sensitive to the degree of population mixing associated with global transmission. Therefore, efforts to restructure secondary school students into modular groups may not be as fruitful as other strategies, such as limiting general mixing of the student population (e.g., in the cafeteria, hallways, common spaces, buses) or cohorting strategies that limit the number of students on campus at one time.

##### B.4 Compliance with isolation upon symptoms

The key to mitigating the risk of large outbreaks is to isolate cases early in order to limit the number of days that any infectious individual is at large and potentially exposing new cases. If individuals self-isolate upon the onset of symptoms, this will help considerably. However, some individuals may not recognize COVID symptoms, or may choose not to comply with a mandated isolation period. Figure B11 shows the effect of the individual compliance rate on the average number of school transmission cases. In the absence of other interventions (blue lines), the rate of self-isolation has a substantial impact on outcomes. In the main text we assume that 20% of individuals self-isolate at the onset of symptoms. This value follows from a finding that professional symptom screening fails to identify nearly half of cases in children with viral loads comparable to symptomatic individuals,[37] and a further assumption that a fraction of individuals that do recognize their symptoms will nevertheless attend school for various reasons. Our results may overestimate the number of school transmission cases if compliance with self-isolation upon symptoms is substantially higher in a given population. In any case, once any proactive mitigation strategy (i.e., testing, cohorting, or vaccination) is introduced, the model outcomes become largely insensitive to the rate of self-isolation compliance. These results suggest that decision makers would be wise to incentivize compliance with symptomatic self-isolation, but they also highlight the fact that coordinated, proactive strategies reduce the risk of outbreaks without relying on individual behavior.

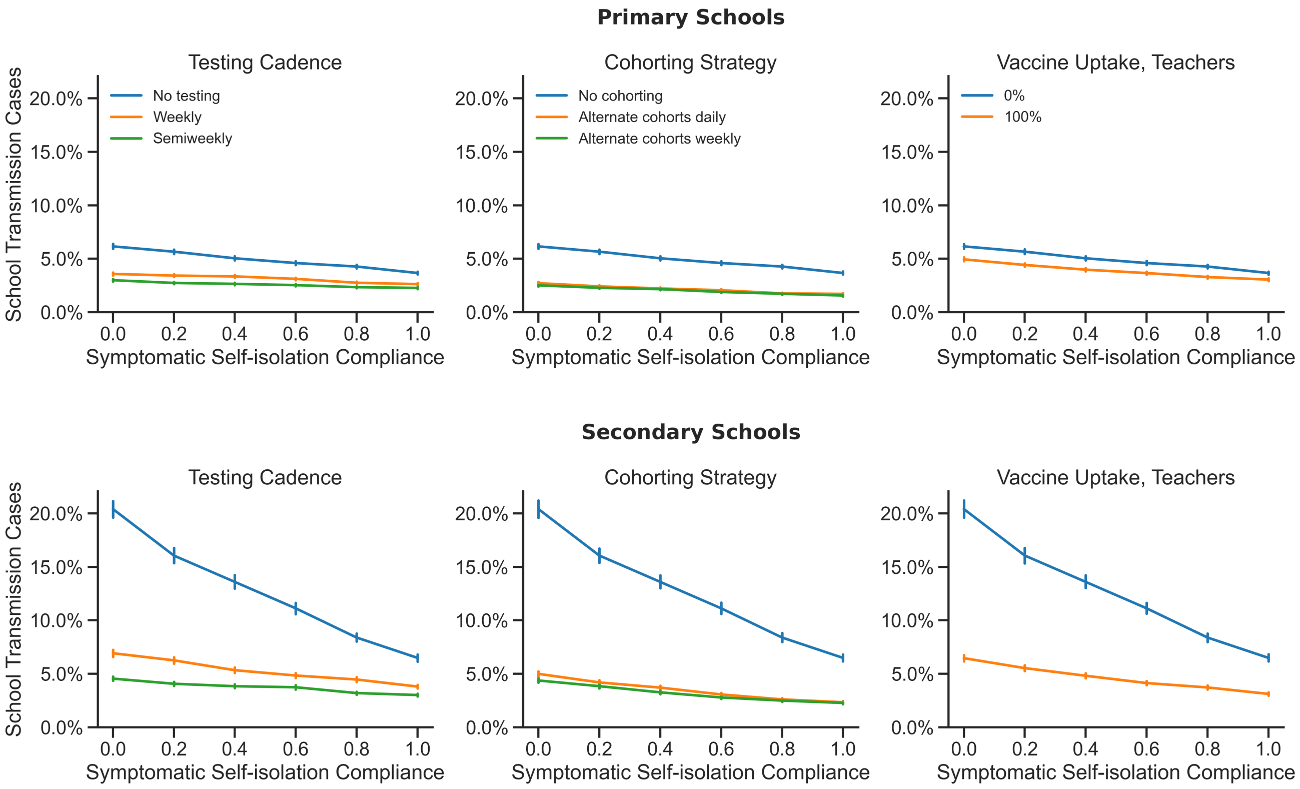

**Figure B11: In the absence of testing, self-isolation compliance is critical to reduce cases, particularly in secondary schools.** Testing detects individuals who are infected but non-compliant, and thereby reduces the importance of the compliance rate. The average number of school transmission cases are shown as a percentage of the total school population for a range of symptomatic self-isolation compliance rates.

#### B.5 Compliance with testing

Our results show that frequent proactive testing is effective at mitigating the risk of sizable outbreaks, but testing is most effective if a large percentage of the population complies with the testing cadence. In the paper we assume that 100% of teachers and staff are tested and that 75% of students comply with testing. We further assume that compliance with testing is a longitudinal property of individuals, rather than determined on a test-by-test basis. Figure B12 shows the effect of varying the rates of student and/teacher compliance with testing on the average number of school transission cases. Unsurprisingly, there are fewer school transmissions on average when a larger percentage of individuals are tested. Outcomes appear to be more sensitive to the level of testing compliance among students than teachers, but this is because students make up a larger fraction of the school population, so drops in student compliance remove a larger absolute number of individuals from the testing pool. The effectiveness of proactive testing is optimized by maximizing the number of individuals that are screened as scheduled. Providing students with adequate incentives to comply with testing forms a necessary component of this strategy.

#### B.6 Test sensitivity (false negative rate)

The success of a proactive testing strategy depends in part on the sensitivity (false negative rate) of the testing methodology used. Test sensitivity changes over time according to disease state of the individual and the amount

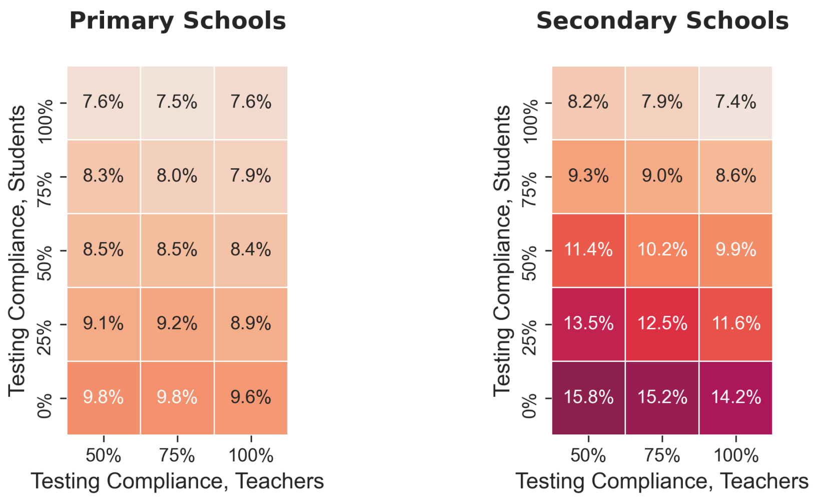

**Figure B12: Compliance with testing by teachers and by students improves outcomes.** Heatmap cells give the average number of school transmission cases. Cases are introduced to the school once a week on average, and baseline values are used for all other parameters as outlined in Table A.2.1.

of time spent in a given disease state (Appendix A.2.6.2). In the main text we base our temporal test sensitivities on data collected for RNA-based tests.[82] Schools may also consider using rapid antigen tests, which are less sensitive than RNA-based tests but are faster and less expensive. Antigen tests have similar peak sensitivities to RNA-based tests around the onset of symptoms, but their sensitivity drops off more quickly after onset and they are less likely to detect pre-symptomatic cases.[96]

Figure B14 compares proactive testing strategies using RNA- and antigen-based testing methods with sensitivity curves drawn from Levine et al. 2020[82] and Smith et al. 2021,[96] respectively (Figure B13). Smith et al. report sensitivities for antigen tests based on their agreement with results from RNA-tests administered at the same time. Here we cap the antigen peak sensitivity values at the corresponding RNA-based sensitivity values for a standardized comparison. Both testing methods significantly reduce school transmission when used weekly or semiweekly. Antigen tests lead to slightly worse outcomes because they are less likely to detect cases during the infectious pre-symptomatic period or late into an individual's infectious period. This difference is more significant in secondary schools where there are generally more cases to detect. In secondary schools, outcomes for weekly RNA-based testing are about equal to those for semiweekly antigen testing.

#### B.7 Out-of-school contacts

When some or all students are out of school, the number of interactions among the school population decreases. If a student is out of school during their infectious period, they are limited in the number of secondary cases they can generate and may fail to transmit disease entirely. Weekends provide a break in exposures between members of the school population, and many other interventions, including cohorting and isolation upon symptoms or positive test, involve keeping students out of school at strategic times. However, if students continue to interact with members of the school population while not in school, then the effectiveness of these exposure breaks will be reduced.

In Figure B15 we consider the effect that out-of-school interactions among students have on the number of transmission cases in the school population. In the main text we assume that a student makes no close contact with

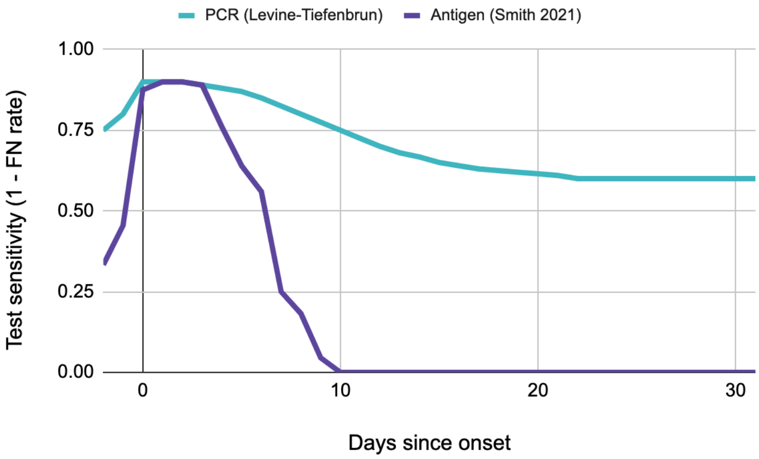

**Figure B13: Test sensitivities vary over the course of infection for RNA-based and antigen methods.** In modeling test sensitivity, we use the following curves for the probability of returning a positive test result for RNA-based tests (teal line, Levine et al. 2020[82]) and antigen tests (purple line, Smith et al. 2021[96]) for infectious individuals as a function of days until or since the onset of the symptomatic or asymptomatic period. The test sensitivity is equivalent to 1 minus the false negative rate.

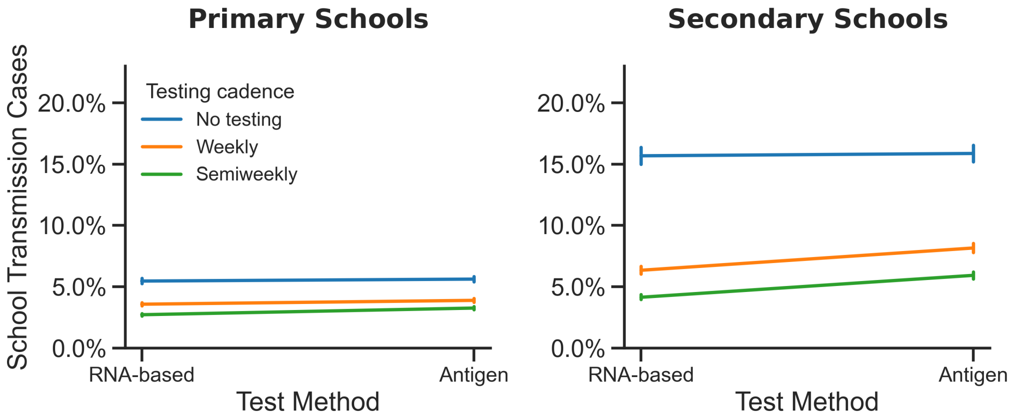

**Figure B14: The benefits of testing are relatively insensitive to testing method**. The average number of school transmission cases are shown as a percentage of the total school population where test sensitivities vary over time according to data corresponding to RNA-based tests[82] or rapid antigen tests.[96] RNA-based tests show a modest advantage in a secondary school setting such that weekly RNA-based testing is comparable to twice-weekly antigen-based testing. This is largely because RNA-based tests are more effective at detecting infection early on during the presymptomatic period. .

any member of the school population (with the exception of any students that share their household) when they are off campus on weekends, cohort off days, or isolation days. Students make incidental contacts with members of the general school population (i.e., global transmission) at the same rate while off campus (e.g., at the grocery store). Here we allow students to maintain some fraction of their close contacts on weekends and on cohort off days where applicable. On off-days, students interact with a fixed random subset of the close contacts they have at school. On weekends, students may maintain contact with any student they interact with in school. Students that are off campus due to cohorting only maintain contact with other students in their cohort. When students continue

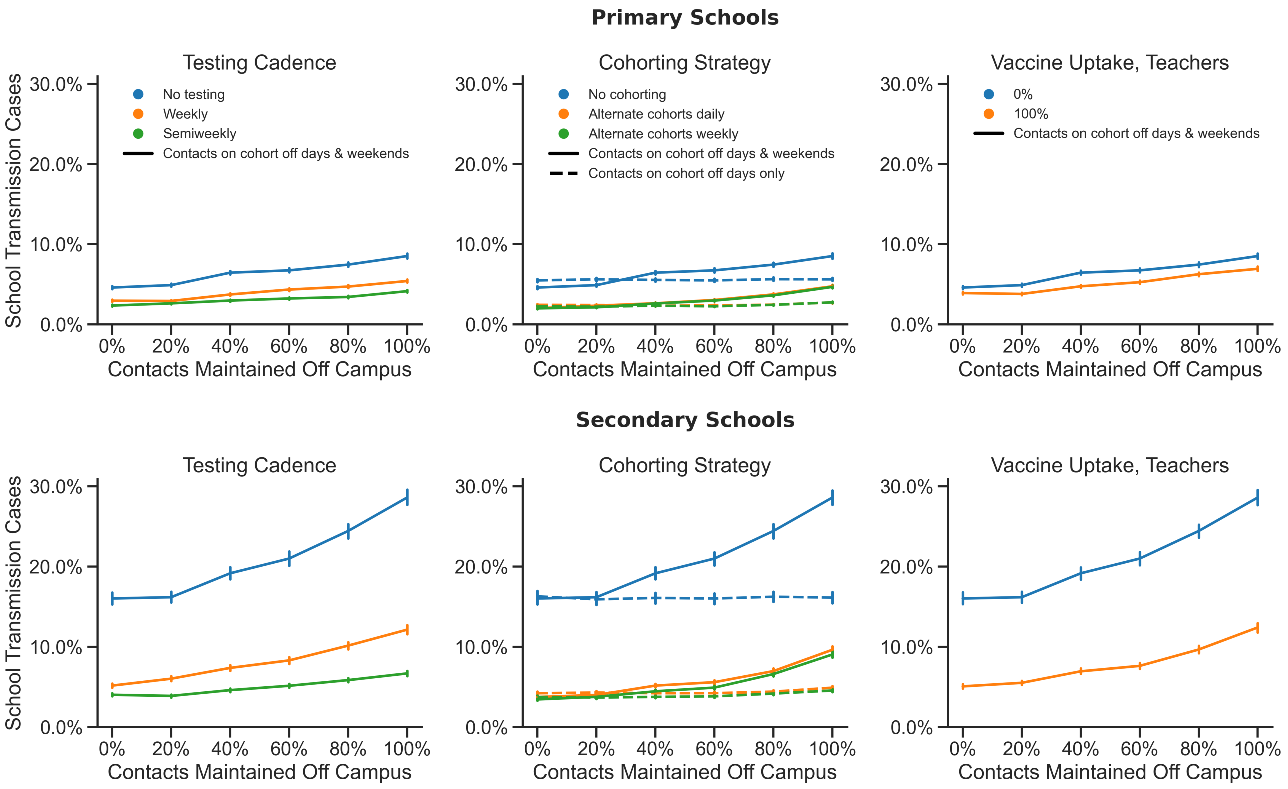

**Figure B15: Weekend interactions among students increase the size of outbreaks.** The average number of school transmission cases are shown as a percentage of the total school population for a range of fractions of school contacts maintained by students while off campus. Outcomes for scenarios where students maintain contacts on weekends as well as cohort off days (where applicable, center column) are shown with solid lines, and outcomes where students maintain contacts on cohort off days only are shown in dashed lines.

to interact at high levels on weekends, the number of school transmission cases increases, which is to be expected given that the total potential exposure time increases by 40%. Interestingly, out-of-school contacts on cohort off days alone do not have a significant effect on the benefit of the cohorting strategy (dashed lines, center column in Figure B15). This suggests that the division of students into two or more groups that never make close contact is sufficient to significantly limit transmission even if these groups continue intra-group interactions while out of school, provided that the cohort boundaries are respected.

#### B.8 Vaccine effectiveness

The success of a vaccination strategy depends on the uptake of the vaccine and its effectiveness in preventing disease and transmission. Uptake and effectiveness have a product relationship, so evaluating a range of values for one parameter or the other gives a good idea of the sensitivity of the model to the effectiveness of a vaccination program in general. We consider the impact of vaccinating all teachers and staff on mitigating transmission in the school population as a whole. Figure B16 shows the influence of vaccine effectiveness on the success of this strategy. Vaccine effectiveness refers to the probability that a vaccinated individual will mount an immune response that consequently blocks disease and infectiousness (Appendix A.2.6.5). Here, we assume that an effective

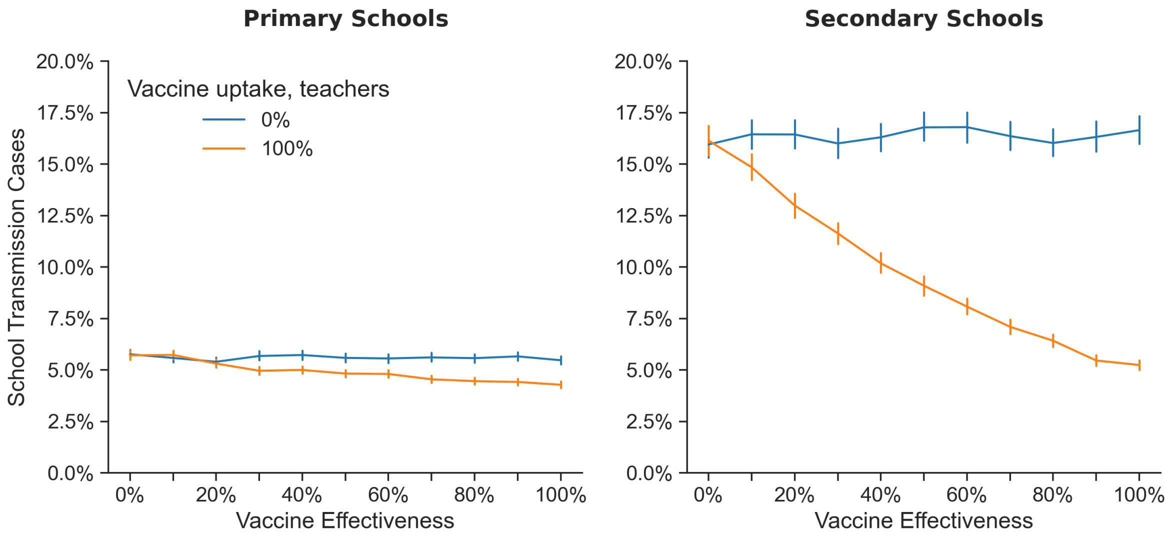

**Figure B16: The benefit of vaccinating teachers in a secondary school setting is sensitive to the efficacy of the vaccine.** The average number of school transmission cases are shown as a percentage of the total school population for a range of vaccine effectiveness values.

vaccination blocks 100% of an individual's propensity for transmission.[62,63] More effective vaccines prevent more school transmission cases on average, but substantive reductions in the mean outbreak size can be achieved even with vaccines that have effectiveness of 50% or lower. Similarly, this suggests that good results can be achieved even with moderate uptake of a highly effective vaccine.

Vaccinating teachers and staff has a greater effect in secondary schools. This is likely because teachers and staff make up a larger fraction of their school populations, interact with more students, and are more likely to transmit to students given the increased susceptibility of secondary school students relative to primary school students.

#### B.9 Classroom isolation

We find that isolating all of an infected student's classroom contacts can mitigate the risk of sizable outbreaks in primary schools (Figure A4), where students are organized into stable classroom units. While this strategy can prevent school transmission, it also requires that additional in-person learning days are lost upon each case detection. When choosing a quarantine strategy, the costs of in-person learning days lost, both by students and by teachers, are likely to be of interest to school decision makers.

Figure B17 shows how both school transmission and in-person learning days missed are impacted by adopting a classroom-level, rather than individual-level, isolation policy. Classroom-level isolation leads to fewer school transmission cases on average, as seen in the negative slope of the lines connecting the points for these two isolation methods for all testing cadences. However, the total number of in-person learning days lost does go up markedly when isolating classrooms rather than individuals.

Figure B17 also shows that when school transmission is relatively low ($R_0 = 1.5$) the marginal reduction in school transmission that is conferred by adopting classroom-level isolation with weekly testing (e.g., moving from the green cross to the green circle) is nearly equal to the reduction that is achieved by switching to semiweekly

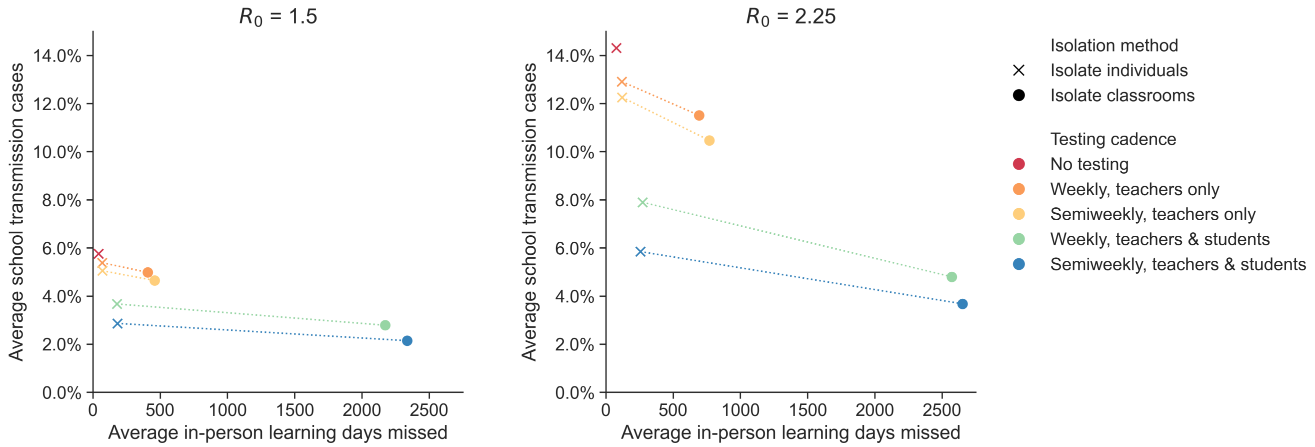

**Figure B17: Isolating entire classrooms can reduce cases, at the expense of missed days of school.** The average number of school transmission cases (as a percentage of the total school population) and the average number of in-person learning days missed (i.e., isolation person-days) are shown for testing and isolation strategies that involve individual-level isolation (crosses) or classroom-level isolation (circles). Results for both isolation policies are shown for five testing cadences, including no testing. A dashed line connects the points for individual-level and classroom-level isolation for each cadence; the slopes of these lines illustrate the relative impact on one outcome variable versus the other. Outcomes are shown for primary schools with weekly introductions and no cohorting or vaccination. Baseline values are used for all other parameters as outlined in Table A.2.1.

testing and isolating only positive individuals (e.g., moving from the green cross to the blue cross). Semiweekly testing leads to a negligible increase in in-person learning days missed over weekly testing, so increasing the testing cadence may be more attractive than adopting a classroom-level isolation policy in some contexts. When transmissibility is higher (right panel, $R_0 = 2.25$), we observe a greater benefit to isolating classrooms, as indicated by the steeper slopes of the lines. .

#### B.10 Cluster Sizes

Throughout this paper, we report the epidemiological outcomes for schools in terms of the total number of school transmission cases and the probability of outbreaks affecting more than 5% of the school population over the course of a semester. These summary statistics convey outcomes that are of primary interest to school decision makers: the expected number of individuals that may become infected at school and the likelihood of an outbreak that causes a major disruption to the health and operations of the school. However, these statistics abstract away more detailed information about transmission chains that can offer insights about the epidemic dynamics. The jitter figures in the main text illustrate the stochastic distribution of school outbreak sizes, and the bimodality of these distributions provides a clue about the underlying dynamics: most transmission chains trains appear either to contribute few to no secondary cases, or alternatively to develop into a large outbreak that may approach the herd immunity threshold if left unchecked.

This observation is confirmed by Figure B18 and Figure B19, which show the distribution of case cluster sizes in primary and secondary schools under different interventions for $R_0 = 1.5$ and $R_0 = 2.25$, respectively. A cluster consists of a primary case that was exposed in the community and any secondary school transmission cases that can be traced back to that primary case. Across both schools and all interventions, a majority of clusters consist of only a single case, which represents an exogenous case introduction that did not generate any secondary cases

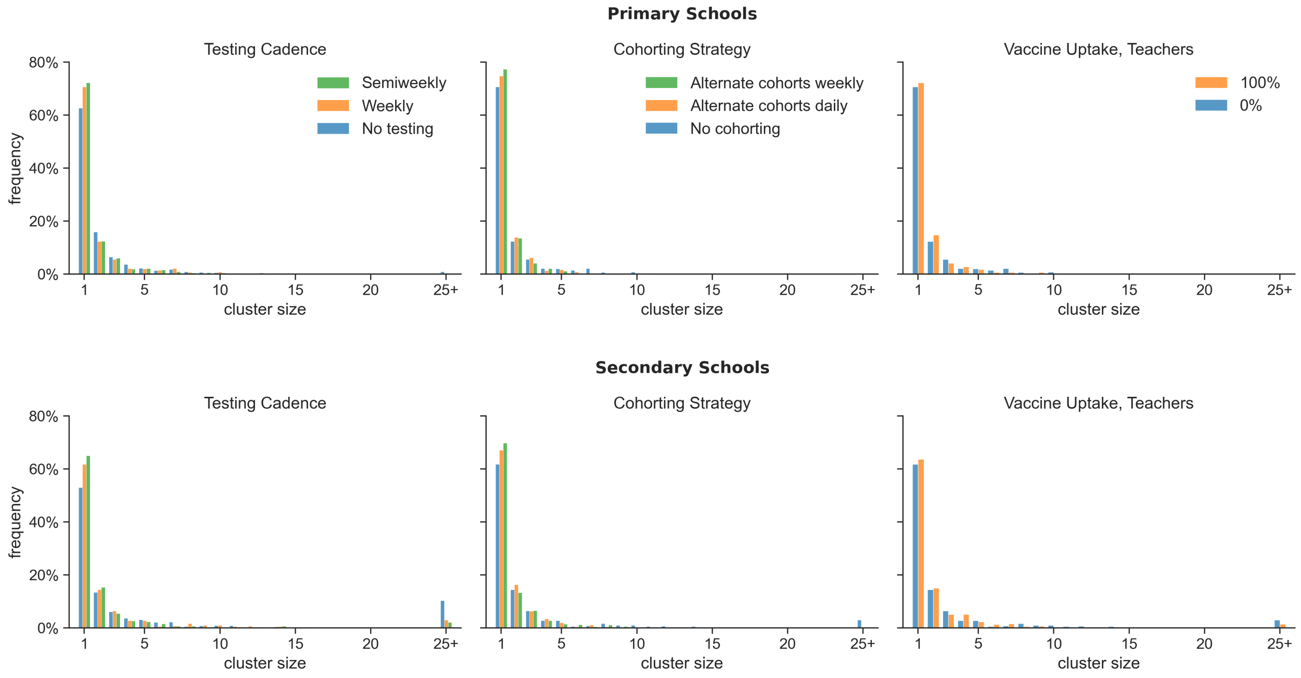

**Figure B18: Distribution of case clusters sizes for $\mathbf{R_0 = 1.5}$.** Histograms give the frequency of case clusters of different sizes in 1000 replicate simulations for each treatment. Data are shown for primary schools (top) and secondary schools (bottom) in scenarios with no mitigation (blue bars), weekly or semiweekly testing of students and teachers (left column), alternating student cohorts on a weekly or daily schedule (center column), and vaccination of teachers (right column). Cases are introduced to the school weekly on average, and baseline values are used for all other parameters as outlined in Table A.2.1.

in the school. In general, smaller clusters are more common than larger clusters. Large clusters (e.g., consisting of 25 individuals or more) are more frequent than intermediate-sized clusters in secondary schools, where school transmission is more likely due to the susceptibility of secondary school students and other factors. When $R_0$ within the school is high (e.g., $R_0 = 2.25$, Figure B19), very large clusters become more common in both settings (which reflects the bimodality seen in the jitter distributions of outcomes for scenarios where $R_0 = 2.25$, e.g., Figure 3 in the main text), and singleton clusters decrease in frequency. Testing, cohorting, and vaccination all increase the frequency of terminal primary cases and decrease the frequency of very large clusters, but these interventions do not dramatically impact the distribution of cluster sizes aside from those extremes.

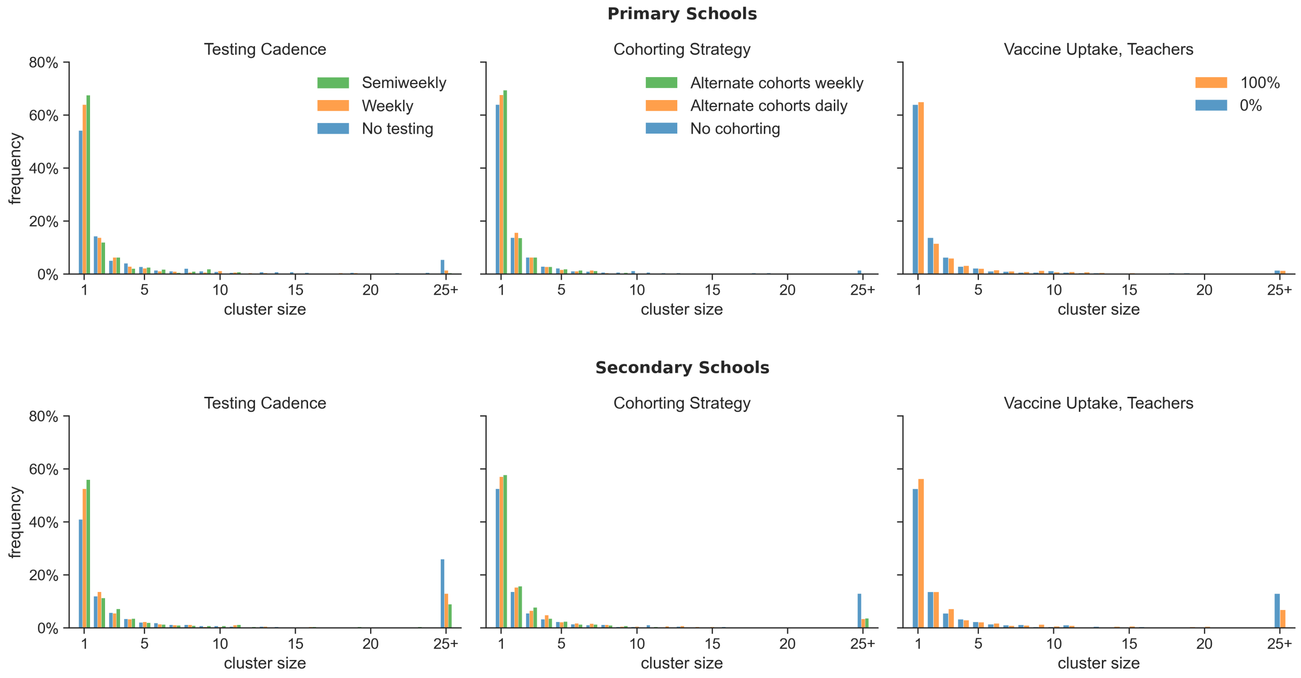

**Figure B19: Distribution of case cluster sizes for $R_0 = 2.25$.** Histograms give the frequency of case clusters of different sizes in 1000 replicate simulations for each treatment. Data are shown for primary schools (top) and secondary schools (bottom) in scenarios with no mitigation (blue bars), weekly or semiweekly testing of students and teachers (left column), alternating student cohorts on a weekly or daily schedule (center column), and vaccination of teachers (right column). Cases are introduced to the school weekly on average, and baseline values are used for all other parameters as outlined in Table A.2.1.
